# Deficient QPRT drives trans-Golgi NAD^+^ hyperinflation and pathological protein secretion in rheumatoid arthritis

**DOI:** 10.1101/2024.10.27.24316032

**Authors:** Lucy Wanjiru Njunge, Kaijing Liu, Chenghe Xiong, Liuqing Chen, Qifei He, Pei Wang, Guan Huang, Yong Li, Peace Osebhue Abhulimen, Wenxiang Cheng, Qiuliyang Yu

## Abstract

Aberrant protein secretion is a central driver of tissue inflammation and destruction in rheumatoid arthritis (RA). While RA fibroblast-like synoviocytes (FLSs) exhibit prominent endoplasmic reticulum (ER) and Golgi, the mechanism underlying their excessive protein secretion is not fully understood. Here, we identified the deficiency of *de novo* NAD^+^ synthesis enzyme, quinolinate phosphoribosyltransferase (QPRT), as a significant abnormality in RA synovium. QPRT loss counterintuitively inflates NAD^+^ in the trans-Golgi network (TGN) while decreasing NAD^+^ in the cytoplasm, ER and cis/medial-Golgi. QPRT knockdown promoted Golgi membrane expansion and epithelial-to-mesenchymal-transition (EMT)-associated secretome production in RA FLSs by suppressing TGN-residing PARP12, resulting in mTORC1-mediated protein translation and Golgi expansion. Furthermore, QPRT gene therapy restored NAD^+^ balance, corrected Golgi dysfunction, reduced cytokine production, and improved RA severity in mouse models. These findings underscore QPRT’s role in coordinating protein secretion and the regulatory dynamics of compartmentalized NAD^+^, proposing QPRT targeting as a therapeutic strategy for inflammatory, secretory and Golgi-related diseases.

## Introduction

Rheumatoid arthritis (RA) is a chronic and debilitating autoimmune disease that not only affects the joints^1^ but also has systemic implications, including increased risks of RA-associated interstitial lung disease (RA-ILD)^2^, cardiovascular disease^3^ and metabolic syndrome^4^. The disease progression involves persistent synovial inflammation (synovitis) and tissue-destructive pannus formation, leading to cartilage and bone damage^1^. Central to RA pathogenesis is the dysregulation of protein secretion, particularly the overproduction of proinflammatory factors such as tumor necrosis factor-alpha (TNF-α) and interleukin-6 (IL-6). The persistent inflammation driven by these secreted proteins further activates various inflammatory pathways, promoting cytokine release and a subsequent vicious cycle of inflammation and joint tissue destruction. Despite significant advances in developing cytokine-targeting biological therapies, such as TNF inhibitors and interleukin blockers, the pathomechanisms underlying the aberrant cytokine production are not well understood, limiting the therapeutic strategies to the blockade of secreted proteins without addressing the root cause. Consequently, these biological therapies demonstrate inefficacy and adverse side effects, thus underscoring the importance of unraveling the molecular mechanisms underlying pathological protein production.

In the inflammatory microenvironment, fibroblast-like synoviocytes (FLSs) undergo an aggressive phenotypic transformation reminiscent of epithelial-mesenchymal transition (EMT), characterized by a hypersecretory phenotype and metabolic reprogramming^5–8^. Activated RA FLSs secrete a myriad of proinflammatory cytokines, chemokines, growth factors and proteases that elicit immune cell infiltration, synovial membrane hyperplasia, subintimal fibrosis, vascularization, and joint tissue destruction ^6,7,9,10^. Despite the remarkable improvements in our understanding of RA FLSs, the pathomechanisms determining RA FLS’s aberrant hypersecretory and tissue-destructive phenotypes are poorly understood. Recent findings highlighted impaired amino acid metabolism as a causative factor in autoimmune inflammation^11,12^. Notably, low circulating tryptophan and elevated kynurenine (Kyn) and quinolinic acid (QA) are consistently associated with RA pathogenesis^11,13,14^. Downstream the Kyn pathway, QA functions as a substrate of quinolinate phosphoribosyltransferase (QPRT), the rate-limiting enzyme in the *de novo* nicotinamide adenine dinucleotide (NAD^+^) synthesis^15^. Meanwhile, the rate-limiting enzyme of the NAD^+^ salvage pathway, nicotinamide phosphoribosyltransferase (NAMPT), has been reported to increase in RA^16,17^. Despite the known roles of dysregulated Kyn pathway and low NAD^+^ levels in RA patients^16^, the expression and role of QPRT in RA pathogenesis are yet to be determined.

Accumulating evidence demonstrates that QPRT is suppressed in various pathologies, including viral infection^18^, kidney injury^19–21^, and infertility^22^. However, the exact mechanism by which QPRT deficiency contributes to the disease remains obscure. Notably, excessive protein production and secretion are reoccurring in most QPRT-deficient pathogenesis^20,22–24^. QPRT suppression reportedly depletes cytoplasmic NAD^+19^. A disrupted subcellular NAD^+^ homeostasis has been shown to induce deficient adenosine diphosphate (ADP)-ribosylation of the endoplasmic reticulum (ER) chaperon BiP, thereby promoting ER expansion and excessive production of TNF in RA T cells^12^. Elsewhere, accumulation of Kyn or decreased ADP ribosylation in cells was implicated in the disease severity^25^ and autophagy inhibition^26,27^ of systemic lupus erythematosus (SLE) by promoting the persistent activation of the amino acid metabolic sensor: mechanistic target of rapamycin complex 1 (mTORC1). mTORc1 is primarily localized on various endomembrane, allowing for specific spatial regulation of its downstream targets, thereby facilitating metabolite-endomembrane communications^28,29^. In particular, the Golgi apparatus (GA)-mTORc1 has been recently implicated in the phosphorylation and Golgi localization of Golgi proteins, thereby modulating Golgi morphology, extracellular vesicles (EV) secretion and protein trafficking^29,30^. While increased mTORc1 is implicated in immune cell infiltration^31^ and RA FLSs aggressiveness^32^, the mechanisms by which mTORc1 contributes to the excessive protein production in RA FLSs remain unclear. In this context, we aim to identify the downstream effector of QPRT deficiency and elucidate how it promotes hyperactive protein production in RA.

In this study, we coupled NAD^+^ biosensors with metabolic profiling and omics studies to explore the role of the QPRT-NAD^+^ axis in RA FLSs pathogenesis. We demonstrate that QPRT is suppressed in RA synovium in humans and mice. Using organelle-specific NAD^+^ biosensors, we demonstrated that QPRT deficiency in RA FLSs decreases global NAD^+^ but counterintuitively inflates NAD^+^ in the trans-Golgi network (TGN) through the suppression of TGN-residing poly(ADP-Ribose) polymerase (PARP) 12. The dysregulated QPRT-PARP12 signaling promotes persistent activation of mTORc1 and increased GRAPS55 phosphorylation, resulting in increased protein translation and Golgi membrane enlargement. The Golgi defects promote enhanced protein production, secretion, and RA FLS invasiveness, thereby converting RA FLSs into hypersecretory pathological cells. We further showed that QPRT deficiency in RA synovium and several carcinomas correlates with the enrichment of EMT, the inflammatory pathway, and mTORc1 signaling. Finally, we utilized AAV-based gene therapy to deliver QPRT in a collagen-induced arthritis (CIA) mice model. The QPRT gene therapy successfully suppressed joint tissue destruction and cytokine production. These findings highlight the importance of subcellular NAD^+^ profiling and underscore the role of QPRT in maintaining Golgi integrity and function. The study further pinpoints the potential of QPRT targeting in fibroblast transformation, autoimmunity, and cancer by regulating the underlying mechanisms contributing to increased protein production.

## Results

### Synovial QPRT is suppressed in RA

To address whether QPRT is associated with RA, we first analyzed publicly available transcriptome data of synovium tissue from RA patients (GSE1919 and GSE89408). QPRT expression in synovium from patients with established RA was significantly lower than that from healthy control and osteoarthritis individuals (Fig. 1a,b, Extended Data Fig. 1a). In contrast, nicotinamide phosphoribosyltransferase (NAMPT), the rate-limiting enzyme in the NAD^+^ salvage pathway, is upregulated in RA synovium (Extended Data Fig.1b,c). Moreover, in the collagen-induced arthritis (CIA) mice model (dataset GSE61140), QPRT was also suppressed compared to healthy control (Fig. 1c). These data reveal that QPRT expression in the synovium is negatively associated with RA pathogenesis. In *de novo* NAD^+^ biosynthesis (Fig. 1d), tryptophan (Trp) is the precursor for kynurenine (Kyn), which is readily produced by the enzyme indoleamine 2,3-dioxygenase (IDO). QPRT then commits the kynurenine pathway to NAD^+^ biosynthesis by converting QA to nicotinic acid mononucleotide (NaMN), a *de novo* NAD^+^ biosynthesis precursor^33^. Further assessment of kynurenine metabolites from RA serum demonstrated a decrease in tryptophan, 3-hdroxyanthranilic acid (3-HAA) and NAD^+^, whereas kynurenine (Kyn), picolinic acid and QA were markedly increased (Fig. 1e-h, Extended Data Fig. 1d,e). Notable, the increased quinolinic acid/tryptophan (QA/T) ratio (Fig. 1i) indicated a defective QPRT activity, further confirming the suppressed QPRT levels in RA patients.

**Fig 1a-c:**
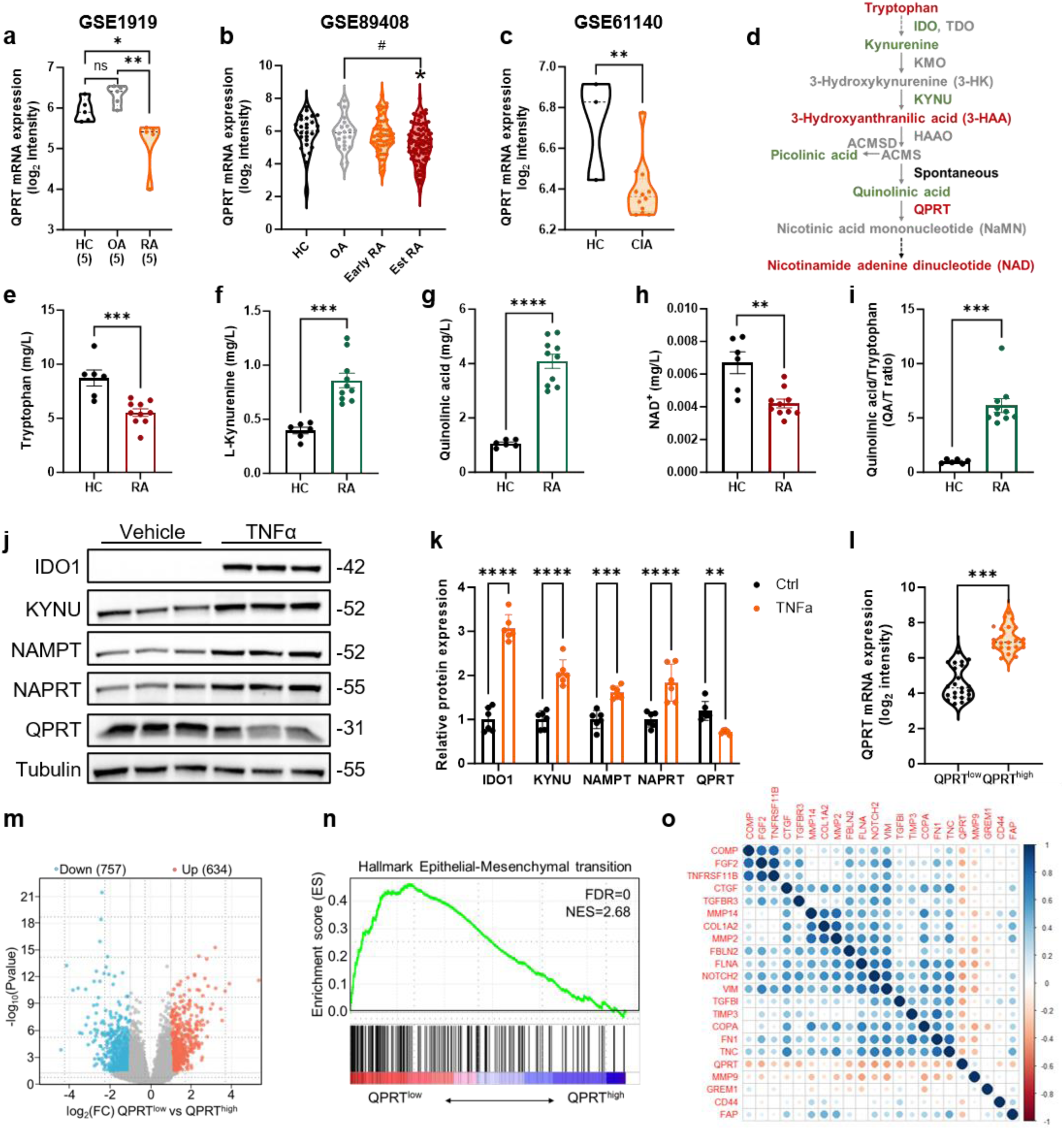
Comparison of QPRT transcripts between healthy controls (HC) and RA human synovium (a-b, GSE1919 and GSE89408) or mouse synovium (c, GSE61140) using publicly available transcriptome data. **d:** Illustration of the kynurenine pathway (KP). **e–h:** LC-MS analysis of KP metabolites from human plasma; HC (n = 6) and RA (n=10) biologically independent samples. **i:** The quinolinic acid (QA) and tryptophan (T) ratio (QA/T ratio) as an indirect representation of QPRT activity **. j:** Representative immunoblot from three independent experiments measuring KP and NAD+ biosynthesis enzyme with and without TNFα (10 ng/mL for 24 hours). **k**: Quantification of KP and NAD^+^ biosynthesis enzyme levels normalized to Tubulin; n = 3 biologically independent samples × n=2 technical replicates. **l:** The top 17% *QPRT* ^high^ expression samples (*n* = 23) and the bottom 17% *QPRT*^low^ expression samples (*n* = 23) from 138 RA patients’ synovial samples pooled from five GEO datasets (GSE12021, GSE55457, GSE55584, GSE77298 and GSE89408). **m:** Volcano plot showing the log_2_fold change (FC) against - log_10_Pvalue comparing from QPRT^low^ and QPRT^high^ RA synovium (n=23 per group). The volcano image was created using the SRplot web server^59^. **n:** GSEA shows the enrichment of the epithelial to mesenchymal transition (EMT) pathway in QPRT^low^ RA synovium. **o:** Correlation coefficient plot of EMT-associated secretome with RA. The correlation coefficients of genes are depicted using a color scheme ranging from red (indicating negative correlation) to blue (indicating positive correlation), with white representing no correlation. Data are mean ± s.e.m.; *P < 0.05, **P < 0.01, ***P < 0.001 and , ****P < 0.0001. ^#^P<0.05 compared to OA. Statistical comparisons were made using one-way ANOVA (**a, b**), two-tailed Student’s t-test (**c,e-i,l**) or two-way ANOVA (**k**).

Fibroblast-like synoviocytes (FLSs) are predominant in the synovial membrane and are key effectors in RA pathogenesis. Using the TNFα-based FLS RA model, we further assessed the effect of inflammation on the major enzymes in the Kyn pathway and NAD^+^ biosynthesis. We showed that QPRT expression alone was negatively regulated by TNFα (Fig.1j,k) among all investigated enzymes involved in the Kyn and NAD^+^ biosynthesis pathway, confirming suppressed synovium QPRT expressions and the involvement of inflammation in regulating QPRT during RA.

We next sought to identify the disease-relevant processes in RA patients with altered QPRT. Analysis of the top 17% synovial samples with the highest (RA QPRT^high^) and lowest QPRT expression (RA QPRT^low^) from 138 RA patients revealed 1391 differentially expressed genes (DEGs), with 634 upregulated DEGs (log_2_ fold change>1, P<0.05) in the QPRT^low^ RA synovium (Fig. 1l,m). Gene-set enrichment analysis (GSEA) defined epithelial-mesenchymal transition (EMT) as the most upregulated signal pathway among all the 50 hallmark gene sets (Fig. 1 n, Extended Data Fig. 1f). We also found that there was a significantly negative correlation between QPRT expression level and several RA hallmarks including TGFβ signaling and angiogenesis (Extended Data Fig. 1f). RA FLSs exhibit an EMT-associated secretory phenotype (EASP)^34^ characterized by the secretion of proinflammatory cytokines, chemokines and matrix metalloproteinases that contribute to RA inflammation, angiogenesis and joint destruction^8^. Data from RA QPRT^low^ and RA QPRT^high^ synovium revealed an inverse correlation between QPRT and several EMT-associated secretome (Fig 1o). Moreover, gene ontology (GO) of biological processes for upregulated genes from the QPRT^low^ group exhibited enrichment of terms involved in EMT, including transmembrane and TGFβ receptor signaling, extracellular matrix remodeling, ossification and wound healing (Extended Data Fig. 1g). Overall, the data implicate QPRT suppression in synovial transformation and EASP during RA.

### QPRT modulates NAD^+^ in trans-Golgi network and Golgi expansion

The intracellular functionality of NAD^+^ signaling depends on a delicate balance between compartmentalized NAD^+^ biosynthesis via the *de novo*, salvage and Preiss-Handler pathways, as well as its consumption by NAD^+^-consuming enzymes (Fig 2a)^35^. To understand the metabolic implication of QPRT deficiency in the different organelles of RA FLSs, we used a compartmentalized NAD^+^ sensor based on bioluminescence resonance energy transfer (BRET) previously developed in our group^36^. The biosensor comprises an NAD^+^-sensitive domain, circularly permuted Nano-luciferase (cpNLuc) and a fluorescent protein mScarlet. Upon NAD^+^ binding to the sensitive domain, a conformational change brings the cpNLuc luciferase and the mScarlet fluorescent protein into proximity (Fig 2b), affecting the intracellular BRET efficiency and hence indicating the NAD^+^ levels of the subcellular environment. For ER targeting, the signaling sequence of calreticulin was incorporated at the N-terminal of the biosensor. Additionally, targeting sequences from monoacylglycerol acyltransferases (MGAT) 2 and galactose-1-phosphate uridylyltransferase (GALT) (Supplementary Table 4) were added to the N terminus to target the biosensor specifically to the cis/medial-Golgi and trans-Golgi, respectively. To quantify NAD^+^ levels in the cytoplasm, ER, cis/medial-Golgi and trans-Golgi, the RA FLSs cell lines, MH7A, were stably transfected with the respective plasmids (Fig 2c).

**Fig 2a:**
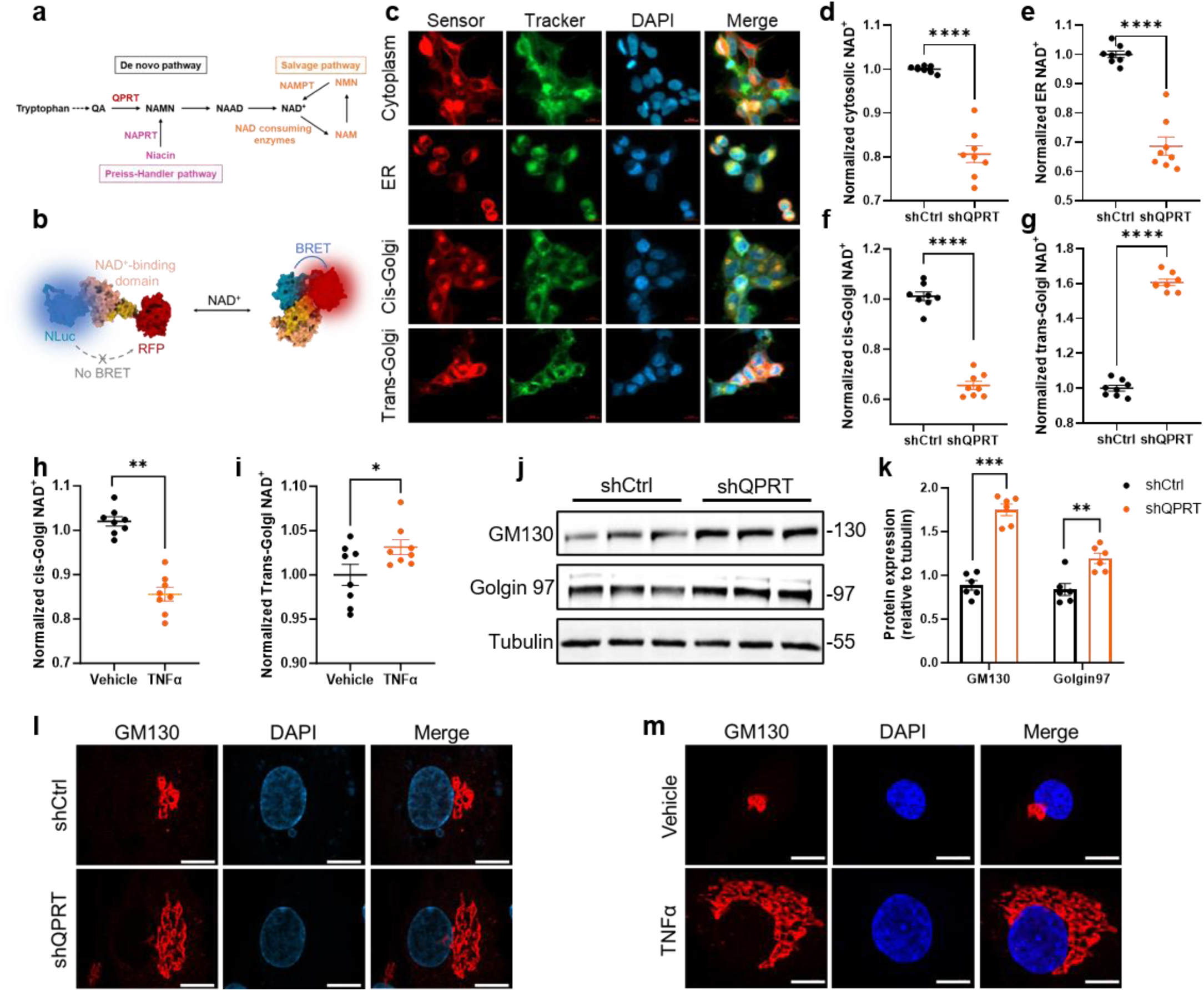
Illustrative diagram of NAD^+^ biosynthesis pathways: *de novo*, salvage and Priess handler pathway. **b:** A schematic representation of the NAD^+^ biosensor structure and mechanism. The sensor comprises cpNLuc, NAD binding domain, and red fluorescent protein (RFP, mScarlet). When NAD^+^ binds, it triggers a conformational change in the sensitive domain, bringing cpNLuc and RFP closer together to facilitate BRET. **c:** Localization of the biosensors (red) in the cytoplasm, ER and Golgi was identified using 488 phalloidin, ER tracker and Golgi trackers (green), respectively. **d-f:** Effect of shQPRT/shCtrl on cytoplasmic, ER, cis/medial-Golgi and trans-Golgi NAD^+^. **h,i:** Effect of TNFα (10ng/ml) on cis/medial-Golgi and trans-Golgi NAD^+^. **j,k:** Representative immunoblot (j) and quantification (k) of the cis/medial-Golgi marker GM130 and the trans-Golgi marker Golgin 97. **l,m:** Representative immunofluorescence image of Golgi marker GM130 in shQPRT/shCtrl transfected RA FLSs (l) and TNFα (10ng/ml) (m). Data are the mean ± s.e.m of independent replicates. P values were determined by two-tailed Student’s t-test (**d-i**) or two-way ANOVA (**k**): *P < 0.05, **P < 0.01, ***P < 0.001 and ****P < 0.0001.

shRNA-based QPRT-knockdown (KD) MH7A cell lines exhibited decreased NAD^+^ levels in the cytoplasm, ER and cis/medial-Golgi network (C/MGN, Fig 2d-f). Unexpectedly, QPRT KD in MH7A induced inflation of NAD^+^ in the trans-Golgi network (TGN, Fig 2g). Accordingly, the changes in NAD^+^ in the C/MGN and TGN were time-dependent (Extended Data 2a-b). Furthermore, TNFα stimulated MH7A cells expressing the organelle-specific NAD^+^ biosensors emulated the QPRT KD phenotype in the cytoplasm, ER, C/MGN and TGN in terms of NAD^+^ modulation (Fig 2h,i, Extended Data Fig. 2c,d). To understand the molecular consequence of QPRT knockdown in the Golgi, we profiled the protein expression of the C/MGN and TGN Golgi proteins. Immunoblotting revealed an increased C/MGN protein GM130 and TGN protein Golgin 97, indicating Golgi expansion (Fig 2j,k). To confirm this observation, we visualized the GA following QPRT knockdown or TNFα activation of human RA FLS. We found that the Golgi was enlarged in both QPRT KD and TNFα activated RA FLSs (Fig 2l,m). Collectively, these data suggest that QPRT differentially regulates the NAD^+^ bioavailability in TGN compared to the cytoplasm, ER and Golgi in the RA FLSs. The inflated NAD^+^ in TGN caused by QPRT loss correlates with the Golgi expansion, highlighting the potential of subcellular NAD^+^ as an indicator of organelle homeostasis and integrity.

### QPRT deficiency enhances RA FLS transformation and secretion

To understand the molecular mechanisms by which QPRT regulates RA FLS function, we profiled the gene expression of the shQPRT and control (shCtrl) RA FLSs by RNA sequencing analysis. Interestingly, most positively regulated genes exhibited a small p-value but minimal fold change, suggesting that the QPRT regulatory mechanism might be at a post-transcriptional level. In agreement with the QPRT^low^ RA synovium data, GSEA defined EMT as the most enriched pathway in QPRT knockdown RA FLSs compared to control (Fig. 3a, b). We further confirmed the role of QPRT in EMT by analyzing the expression of EMT markers and invasiveness in QPRT knockdown RA FLSs. Consistently, QPRT deficiency induced the expression of ZEB1, SNAIL, PDPN and N-cadherin (CDH2) and increased RA FLSs invasiveness compared to control (Fig. 3c,d Extended Data 3a). Notably, GO analysis showed an overrepresentation of secretory pathway-related terms including ER to Golgi vesicle-mediated transport, Golgi vesicle transport, ER lumen and ER-Golgi intermediate compartment (Fig. 3e). Moreover, there was an enrichment of terms corresponding to extracellular matrix organization, focal adhesion, cell-substrate adhesion, collagen-containing extracellular matrix, wound healing, cell-substrate adhesion and extracellular matrix organization (Fig. 3e). In agreement with previous reports on QPRT’s proliferative role, the knockdown of QPRT markedly suppressed genes related to cell cycle and cell division (Extended Fig. 3b). Although multiple factors could modulate RA FLS transformation, we found that QPRT negatively correlated with EMT-associated secretome including growth factors, cytokines, ECM components, chemokines and proteases (Fig. 3f-h, Extended Data Fig 3c). Overall, the data suggest that QPRT restrains RA FLSs transformation and loss of QPRT promotes the ER-Golgi secretory pathway and the production of EMT-associated secretome.

**Fig 3a:**
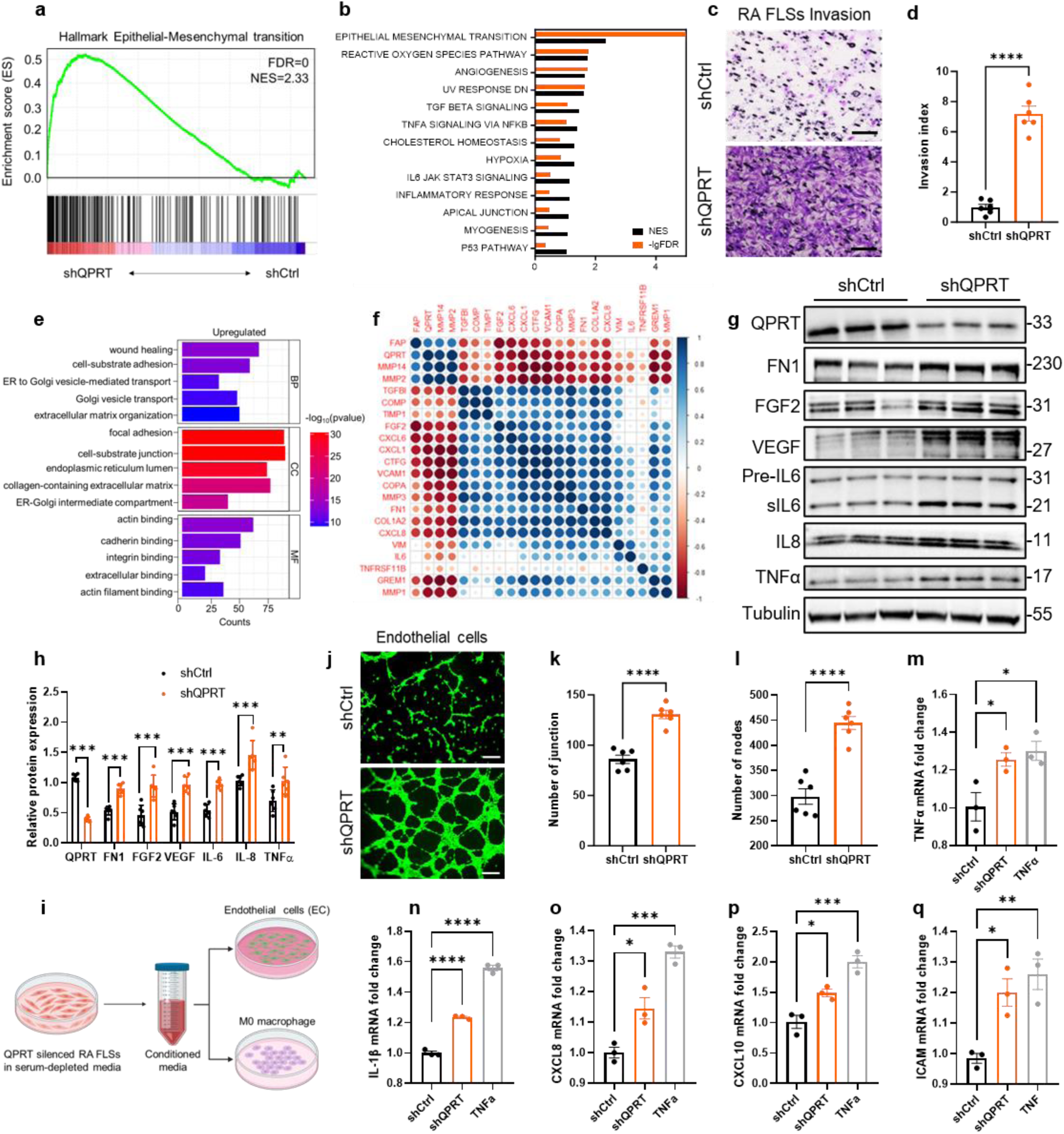
GSEA shows the enrichment of the epithelial to mesenchymal transition (EMT) pathway in shQPRT/shCtrl transfected RA FLSs. **b:** Normalized Enrichment Scores (NES) with −log_10_FDR for significant pathways identified in the GSEA analysis are presented. The FDR is zero for the EMT pathway, resulting in a −log_10_FDR of infinity. Detailed data can be found in Supplementary Table 2. **c:** Representative images of Transwell invasion assay. **d:** Quantification of relative invasive rate (relative to shCtrl group). **e:** Gene ontology (GO) analysis on positively regulated genes. BP, biological properties; CC, cellular components; MF, molecular function. GO bar plot was created using the SRplot web server^59^. **f:** Correlation coefficient plot of RA-associated EMT-related genes in shQPRT/shCtrl transfected RA FLSs. The correlation coefficients of genes are depicted using a color scheme ranging from red (indicating negative correlation) to blue (indicating positive correlation), with white representing no correlation. FDR, false discovery rate; NES, normalized enrichment score. **g,h:** Representative immunoblotting images (f) and quantification (g) of EMT-associated secretome. **i:** Schematic representation of the coculture of endothelial cell line (EA.hy926) and macrophage cell line (THP1) with conditioned medium from shQPRT/shCtrl transfected RA FLSs. **j:** Capillary tube formation of EA.hy926 cultured in condition medium from shQPRT/shCtrl transfected RA FLSs. **k,l:** Quantification of the number of junctions and nodes analyzed by Angiogenesis Analyzer plugin on image J. **m-q:** mRNA expression levels of M1 macrophage markers TNFα, IL-1β, CXCL8, CXCL10 and ICAM in THP1 cells cultures in CM derived from shQPRT/shCtrl transfected RA FLSs. Data are the mean ± s.e.m of independent biological samples. P values were determined by two-tailed Student’s t-test (**d,k,l**), two-way ANOVA (**h**) or one-way ANOVA (**m-q**): *P < 0.05, **P < 0.01, ***P < 0.001 and ****P < 0.0001.

Based on the above observation that QPRT loss led to increased protein production of secretory factors related to RA pathogenesis, we sought to elucidate the biological activity of the secreted factors in RA FLS communication with macrophage and endothelial cells. Upon coculture with conditioned media obtained from QPRT knockdown or Ctrl RA FLSs, tube formation was highly enhanced in endothelial cells, whereas M1 macrophage markers, not M2 markers, were induced by the knockdown media compared to the Ctrl media (Figi-q, Extended Data Fig 3d-g). Accordingly, ELISA assay and western blot of secreted proteins detected increased secretion of inflammatory and angiogenic factors (Extended Data Fig. 3h-k). Therefore, increased ER-Golgi vesicle trafficking might increase protein secretion in QPRT-perturbed RA FLSs, resulting in increased crosstalk with other joint cells propagating disease progression.

### PARP12 loss accounts for QPRT-knockdown phenotypes

As QPRT loss led to inflated TGN-NAD^+^, expanded Golgi, and upregulated ER-Golgi vesicle trafficking and protein secretion, we sought to identify the effector protein of QPRT deficiency in the Golgi. We first performed proteomics analysis in control and QPRT knockdown cells. A comprehensive protein expression analysis revealed 138 differentially regulated proteins (0.5 < log2 fold change < –0.5, P < 0.05) after QPRT knockdown (Fig. 4a, b). GO analysis of upregulated terms showed a substantial overrepresentation corresponding to ribosome biogenesis (e.g., RPSA, RPS13, RPS17, RPS16), nucleocytoplasmic transport, and cytoplasmic translation (Fig. 4c), suggesting that QPRT might influence protein production by modulating the protein translation machinery. Moreover, functional analysis of downregulated proteins revealed strong enrichment of proteins involved in nucleotidyltransferase activity, NAD^+^-protein ADP-ribosyltransferase activity and glycosyltransferase activity (Fig. 4d).

**Fig 4a:**
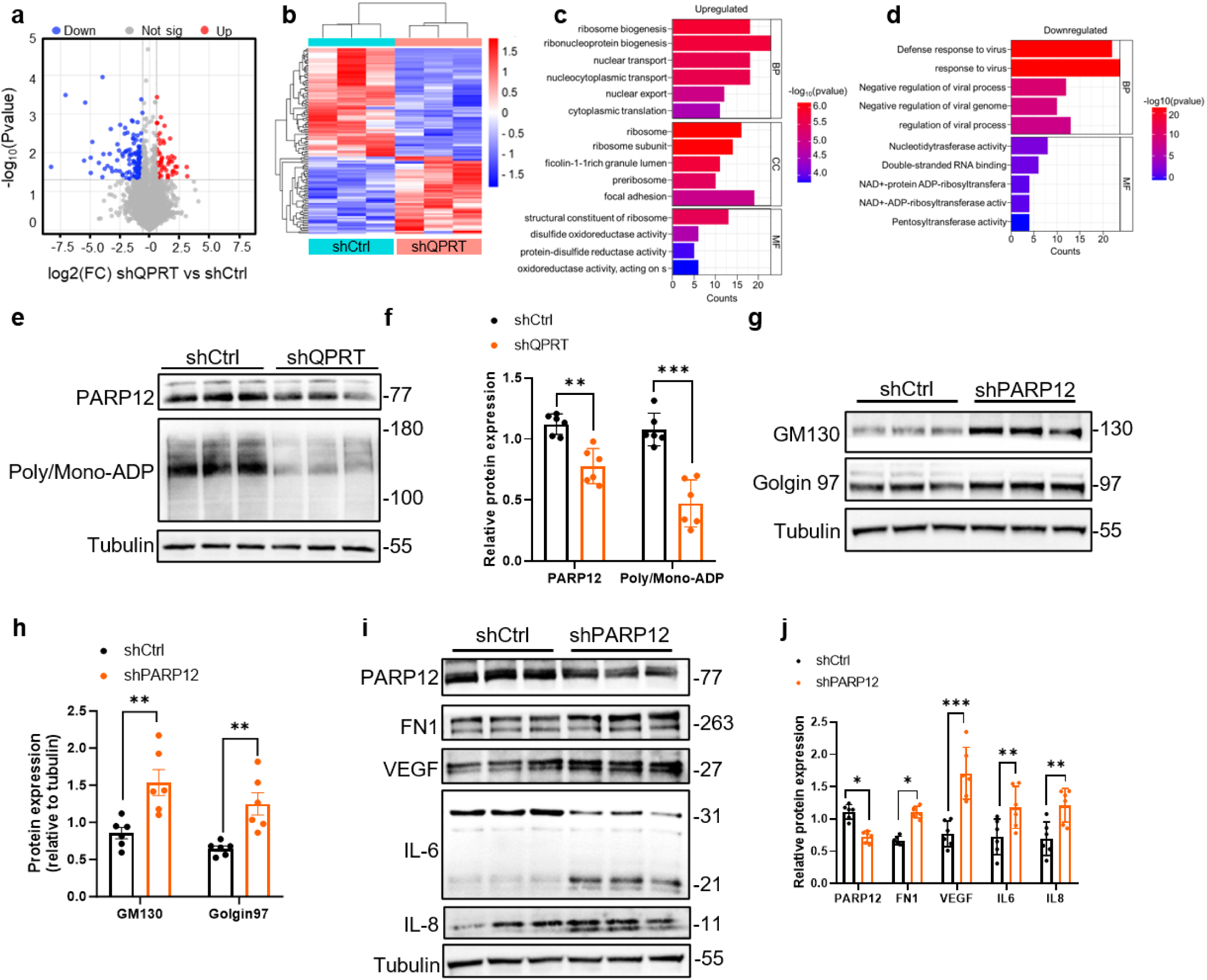
Volcano plot showing the log2fold change (FC) against -log10Pvalue comparing from shQPRT/shCtrl transfected RA FLSs (n=3 per group, P < 0.05, |logFC|>1.0). **b:** Heatmap plot of differentially expressed proteins. The complete cluster algorithm and Euclidean distance metric were used. **c,d:** Gene ontology (GO) analysis on upregulated (c) and downregulated (d) proteins. BP, biological properties; CC, cellular components; MF, molecular function. **e,f:** Representative immunoblot (e) and quantification (f) of PARP12 and Poly/Mono-ADP ribose levels. Representative and collective data from three biological and two technical replicates. **g,h:** Representative immunoblot (g) and quantification (h) of the cis-Golgi marker GM130 and the trans-Golgi marker Golgin 97 in shPARP12/shCtrl transfected RA FLSs**. i,j:** Representative immunoblotting images (i) and quantification (i) of EMT-related secretome in shPARP12/shCtrl transfected RA FLSs. Data are the means ± s.e.m. Statistical comparisons were made using two-way ANOVA: \**P* < 0.05, \*\**P* < 0.01, \*\*\**P* < 0.001. The volcano plot (**a**), heatmap (**b**) and GO bar plot(**c,d**) were created using the SRplot web server^59^.

Considering the distinct NAD^+^ inflation in TGN, we focused on proteins related to the Golgi. Trans-Golgi residing mono-adenosine diphosphate (mADP)-ribosyltransferase, PARP12, was among the downregulated ADP-ribosyltransferase implicated in protein translation, Golgi integrity and secretion. Western blot further verified PARP12 and ADP downregulation in QPRT knockdown RA FLSs (Fig 4e, f). At the same time, the confocal microscope showed a decrease of perinuclear localized PARP12 in QPRT knockdown cells (Extended Fig. 4b). When PARP12 was knocked down in RA FLS, we observed that PARP12 knockdown phenocopied QPRT knockdown in relation to Golgi expansion (Fig. 4g,h) and RA-related secretome (Fig. 4i,j). Collectively, these data identify PARP12 as a QPRT-regulated protein that modulates Golgi size and function and RA FLSs secretory phenotype.

### PARP12 facilitates GRASP55 dephosphorylation

Having established that the QPRT-PARP12 signaling modulates Golgi expansion, protein production and secretion, we next sought to elucidate the underlying mechanisms. mTORc1 is a central regulator of protein translation through the phosphorylation of ribosomal protein S6 kinase 1 (referred to here as S6) and the eukaryotic initiation factor 4E binding protein 1 (4E-BP1), thereby promoting ribosome biogenesis and protein translation, respectively^37,38^. Furthermore, mTORc1 was recently shown to phosphorylate the TGN-residing stacking GRASP55, resulting in its Golgi retention^30^. Given the essential role of mTORc1 in protein production and GRASP55 phosphorylation, we sought to examine whether the QPRT-PARP12 axis modulated mTORc1 activity in RA FLSs. Phosphorylation of GRASP55 and S6 was dramatically enhanced in QPRT and PARP12 knockdown RA FLSs, confirming the activation of mTORc1 in these cells (Fig 5a-d). The increased phosphorylation of S6 in QPRT knockdown RA FLSs might explain the enhanced ribosomal biogenesis observed in these cells. Consistent with the known mTORc1-specific phosphorylation of GRASP55^30^, QPRT knockdown did not induce the phosphorylation of GRASP65 (Extended Data Fig 5a). Phosphorylation of GRASP55 is known to disrupt its oligomerization, leading to Golgi cisternae unstacking^39^, which has been shown to enhance vesicle formation, protein trafficking and secretion^40^. Consistently, immunostaining of GRASP55 further illustrated Golgi ribbon unlinking, resulting in an enlarged Golgi (Extended Data Fig 5b). Moreover, QPRT suppression induced decreased expression of 4EBP1 (Fig 5a,e). The suppressed 4EBP1 and increased mTORc1 activity promote enhanced protein synthesis by relieving 4EBP1 inhibition of the cap-dependent translation, further highlighting the mechanism by which QPRT suppression increases protein production. Together, these results suggest that the increase in Golgi size and increased protein production and secretion observed in QPRT and PARP12 knockdown RA FLSs might result from mTORc1 activity and the regulation of the translation machinery.

Given that GRASP55 and PARP12 are located in the TGN, we were curious whether these proteins interact. Co-immunoprecipitation showed that PARP12 interacts with GRASP55, and the knockdown of QPRT diminished the interaction (Fig. 5g). To assess whether QPRT or PARP12 loss impacted GRASP55 ADP-ribosylation, GRASP55 protein was immunoprecipitated from QPRT knockdown, PARP12 knockdown, and Ctrl RA FLSs. As shown in Fig. 5h, QPRT or PARP12-deficient RA FLSs contained deribosylated GRASP55, indicating that the QPRT-PARP12 axis promoted GRASP55 ribosylation. Moreover, overexpression of PARP12 and GRASP55 in the presence of NAD^+^ markedly increased GRASP55 ADP-ribosylation (Fig 5i), further confirming the ribosylation capacity of PARP12.

**Fig 5a,b:**
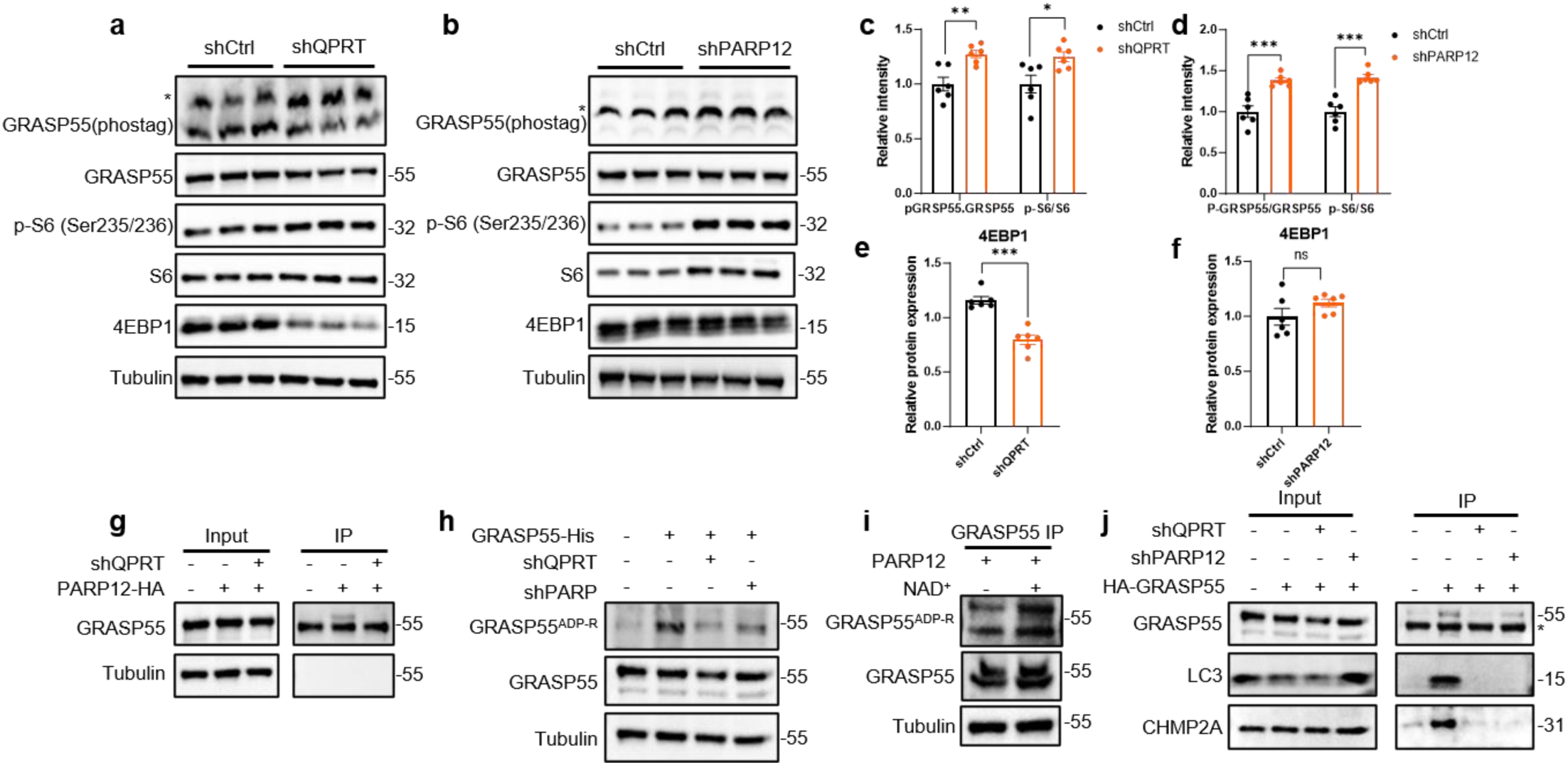
Immunoblotting analysis was performed on lysate from shCtrl, shQPRT (a), or shPARP12 (b) transfected RA FLSs. GRASP55 phosphorylation was assessed using phos-tag gels, and the asterisk indicates p-GRASP55. mTORc1 activity was assessed by the phosphorylation of ribosomal protein S6 (S6) and eukaryotic initiation factor 4E-binding protein 1(4E-BP1). **c-f:** Quantification of the proteins. **g:** Co-immunoprecipitation and immunoblotting experiments confirm PARP12 interaction with GRASP55. **h:** ADP ribosylation of GRASP55 in shQPRT or shPARP12 transfected RA FLSs. **i:** ADP ribosylation of GRASP55 in PARP12 overexpressing RA FLSs in the absence or presence of 100 μM NAD+ for 24 h (n = 3). **j:** Co-IP analysis of GRASP55 interaction with the autophagosome marker LC3B, and multivesicular body (MVB) marker CHMP2A in shCtrl, shQPRT and shPARP12 transfected RA FLSs. Data are the means ± s.e.m. Statistical significance was assessed by two-way ANOVA (**c,d**) or two-tailed Student’s t-test (**e,f**): ns, not significant, \**P* < 0.05, \*\**P* < 0.01, \*\*\**P* < 0.001.

Dephosphorylated GRASP55 is proposed to promote autophagy-dependent unconventional protein secretion (UPS) by relocating from the Golgi to autophagosome and multivesicular bodies (MVB)^30,41^. Consistently, we found that GRASP55 binding to autophagosomal marker LC3B and MVB marker CHMP2A was significantly reduced by QPRT and PARP12 knockdown (Fig 5j), further implicating the QPRT-PARP12 axis in the regulation of GRASP55-dependent UPS. Interestingly, QPRT knockdown decreased LC3B without affecting p62 expression, indicating autophagic blockage (Extended Data 5c,d). Moreover, the knockdown of PARP12 induced the accumulation of LC3B and p62, suggesting impaired autophagosome-lysosome fusion (Extended Data Fig 5e,f). The effect of PARP12 depletion on autophagosome maturation is similar to that reported in GRASP55 knockdown cells^41^, confirming the interaction between these two proteins. The dysregulated autophagy and disrupted interaction of GRASP55 with autophagosome and MVB components in QPRT- or PARP12-depleted RA FLSs suggest that the QPRT-PARP12 axis might be essential in regulating autophagy and GRASP55-dependent unconventional protein secretion.

Together, these data define PARP12 and NAD^+^-dependent ADP ribosylation of GRASP55 as an on-off switch of Golgi-dependent and autophagosome-dependent GRASP55 signaling, placing QPRT upstream of Golgi morphology and function, mTORc1 activation and autophagy.

### Therapeutic and prophylactic efficacies of AAV-QPRT

Given that QPRT is consistently downregulated in RA synovium, and its deficiency in cells leads to increased protein production and secretion, we sought to determine whether AAV2-QPRT gene therapy would improve RA outcome in a murine model of collagen-induced arthritis (CIA) with empty AAV2 vectors (Vector) as control (Fig 6a). At the endpoint, we confirmed that QPRT was overexpressed in mice paws (Extended Data Fig 6a,b). We further confirmed that overexpression of QPRT induced NAD^+^ elevation in AAV-QPRT mice compared to the Vector group (Extended Data Fig 6c). Clinical and morphological assessment revealed that the arthritis score and paw size of the AAV-QPRT group were significantly smaller compared to the Vector group, indicating decreased disease activity and severity (Fig 6b-c). Although QPRT effectively mitigates symptoms and disease severity, overexpressing QPRT in established RA is insufficient to prevent disease onset (Fig 6d). Hematoxylin and eosin (H&E)- and Safranin O-based histopathological analysis demonstrated minimum degradation of the articular cartilage and bone (Fig.6e, Extended Data 6d). Moreover, tartrate-resistant acid phosphatase (TRAP) confirmed that QPRT protected bone resorption by inhibiting osteoclast activity during RA, as demonstrated by reduced TRAP+ preosteoclasts and osteoclasts in AAV-QPRT mice (Fig. 6e). In contrast, the cartilage and bone tissue from the Vector group was severely damaged, with an intense ectopic lymphoid structure (ELS) observable in the joints (Fig.6e). In addition, micro-computed tomography (micro-CT) analysis was performed to assess bone erosion. As shown in Fig f-i, quantitative bone mineral density (BMD), trabecular thickness, and percentage bone volume analysis demonstrated that the QPRT overexpression preserved bone integrity. Altogether, these data suggest that overexpression of QPRT in RA protects joint tissue from tissue-destructive activities of RA.

**Fig 6a:**
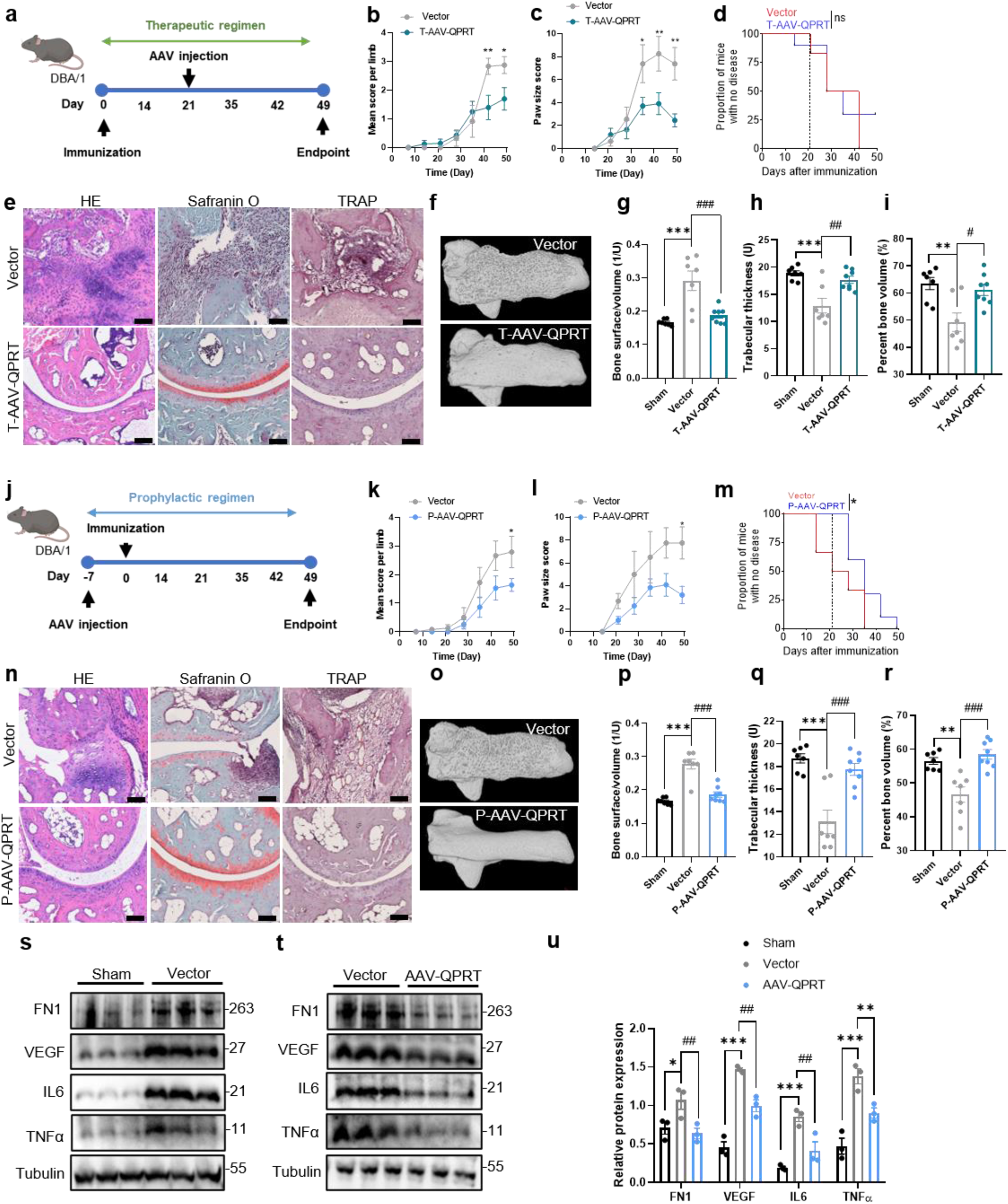
Schematic illustration of the therapeutic (T) regimen. Following immunization with type II collagen, mice were randomly assigned to three groups (sham and Vector n=7 /group, AAV-QPRT, n=8). On day 21, mice either received a single injection of rAAV-CMV-WPRE-pA; AAV2/2 (Vector) or rAAV-CMV-Qprt(m)-WPRE-pA; AAV2/2 (AAV-QPRT). **b,c:** Disease severity and activity were assessed using arthritis scores (b) and hind paw thickness (c). **d: D**isease prevalence was measured from day 21 after immunization once a week. **e:** Representative histology images of hematoxylin-eosin (H&E, left), safranin O/Fast green (middle) and TRAP staining (right) of the ankle joint from Vector and AAV -QPRT mice. **f:** Micro-CT representative image of the calcaneus bone. **g-i:** Bone surface-to-volume ratio (BS/BV) (g), trabecular thickness (Tb Th) (h), and percentage bone volume (i) in the calcaneus bone were assessed by micro-CT and 3D reconstruction. **j:** Schematic illustration of the prophylactic (P) regimen. Mice were randomly assigned to two groups (Vector n=7, AAV-QPRT, n=8) and injected with a single dose of rAAV-CMV-WPRE-pA; AAV2/2 (Vector) or rAAV-CMV-Qprt(m)-WPRE-pA; AAV2/2 (AAV-QPRT) seven days before immunization. **k-m:** Following CIA induction, arthritis scores (k), hind paw thickness (l) and disease prevalence (m) were measured once a week. **n:** Representative H&E (left), Safranin O/Fast green (middle) and TRAP staining (right) of the hind ankle joint from Vector and AAV-QPRT mice. **o:** Micro-CT images of the calcaneus bone**. p-r:** BS/BV (p), Tb Th (q), and BV/TV (r) in the calcaneus bone were assessed by micro-CT and 3D reconstruction. **s,t:** Hind paws from sham, Vector and AAV-QPRT were homogenized in cell lysis buffer for immunoblot detection of FN1, VEGF, IL-6 and TNFα. **u:** Quantification of FN1, VEGF, IL-6 and TNFα levels in sham, Vector and AAV-QPRT mice. Immunoblot represents data pooled from eight mice per group. Statistical comparisons were made using one -way ANOVA (**g-i, p-r**), Log-rank (Mantel-Cox) test (**d, m**) or two-way ANOVA (**u**). ns, not significant, *P < 0.05, **P < 0.01, ***P < 0.001 compared to sham, ^##^P < 0.01 compared to Vector.

To evaluate the broader application of AAV-QPRT, we tested its prophylactic regimen (Fig 6j). Paw sizes measured from mice in the AAV-QPRT group were significantly smaller than in the AAV-Empty group (Fig 6k,l). Interestingly, overexpression of QPRT delayed the onset and progression rate of the disease compared to the Vector group (Fig 6m), suggesting that QPRT might play an essential role in blocking the proinflammatory acquisition during RA development. Histological analysis also confirmed a reduction of immune infiltration and preservation of cartilage and bone in mice receiving AAV-QPRT (Fig 6n). The micro-CT and TRAP staining indicated that arthritis-associated bone erosion was significantly reduced in the AAV-QPRT mice compared to the Vector group (Fig 6o-r). Therefore, targeting QPRT during the early stages of RA development might prove advantageous in preventing disease incidence and severity.

Collectively, these data underscore the therapeutic potential of targeting QPRT in RA treatment. Moreover, the prophylactic targeting of QPRT yields superior outcomes compared to the overexpression of QPRT after RA is established. By overexpressing QPRT before the onset or during the early stages of RA, QPRT could effectively revert inflammatory processes that lead to joint damage and disease progression, thereby reducing the severity of symptoms and risk of irreversible joint deformation. Mechanistically, QPRT overexpression effectively blocked the production of proinflammatory factors in the joints, as demonstrated by the suppression of VEGF, IL6 and TNFα in the AAV-QPRT mice compared to the Vector group (Fig 6s-u). Consistently, RA induced the decreased expression of PARP12 and increased expression of GRASP55, which were reversed by the overexpression of QPRT (Extended Data Fig 6f-h).

### QPRT expression and EMT in cancer

In addition to the previously reported suppression of QPRT in kidney renal clear cell carcinoma, kidney chromophobe and kidney renal papillary cell carcinoma (Fig. 7a-c)^42^, we also identified prostate adenocarcinoma (PRAD), and cervical squamous cell carcinoma and endocervical adenocarcinoma (CESC) as QPRT suppressed carcinomas as demonstrated by the cancer genome atlas (TCGA) datasets(Fig 7d,e). To explore the potential mechanisms by which QPRT suppression promotes renal cancer, we utilized the TCGA KIRC dataset (TCGA-KIRC), including 541 clear cell renal cell carcinoma (ccRCC) patient samples. The GSEA analysis of 10% highest QPRT (QPRT^high^) and 10% lowest QPRT (QPRT^low^) mRNA expression (54 samples per group)^20^ revealed a significant enrichment of TGFβ, ultraviolet (UV) response, mitotic spindle and EMT signaling pathways in QPRT^low^ samples (Extended Data Fig7a,b). Analysis of the clinical proteomic tumor analysis consortium (CPTAC) datasets further identified head and neck squamous cell carcinoma (HNSC), lung squamous cell carcinoma (LSCC), uterine corpus endometrial carcinoma (UCEC), pancreatic ductal adenocarcinoma (PDAC) (Fig 7f-j) and hepatocellular carcinoma (HCC) (results not shown). PARP12 deficiency^43^ and suppression of Kyn enzymes upstream QPRT^44^ in HCC were proposed to promote metastasis by regulating EMT and DNA damage-mediated tumorigenesis, respectively. Consistently, the GSEA analysis of top 10% QPRT^low^ and top 10% QPRT^high^ HCC samples from the TCGA LIHC dataset (TCGA-LIHC, including 371 HCC patient samples), showed a remarkable negative correlation between QPRT expression and EMT, TNFA signaling via NFKB, inflammatory responses, TGFβ and angiogenesis signaling pathways (Fig. 7k, Extended Data Fig7c). Further analysis of enriched pathways in the identified QPRT-suppressed cancers showed an inverse correlation between QPRT expression and various pathways, including the P53 pathway, interferon response, MYC targets, EF2 targets, mTORc1 signaling and protein secretion (Fig. 7l). Together, these results suggest that the QPRT mechanism is likely to be shared across different biological and pathological contexts, extending the potential indications of targeting QPRT beyond RA.

**Fig 7a-e:**
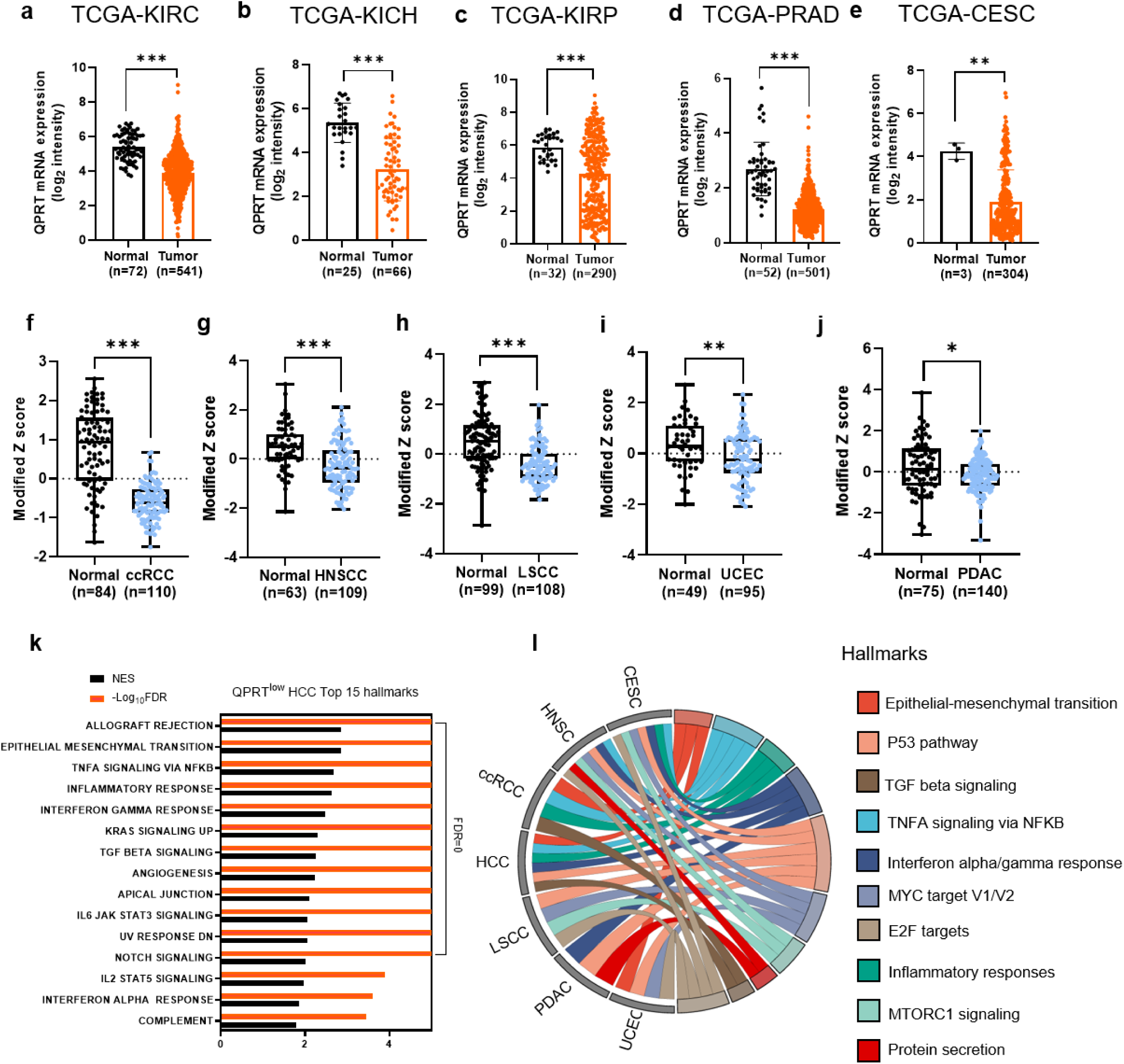
The Cancer Genome Atlas (TCGA) datasets expressing suppressed QPRT transcript. **f-j**: Clinical Proteomic Tumor Analysis Consortium (CPTAC) datasets expressing suppressed QPRT protein. **k:** Normalized Enrichment Scores (NES) with −log_10_FDR for QPRT^low^ (TCGA-LIHC, n=37) significant pathways identified in the GSEA analysis. The FDR is zero for the top twelve pathways, resulting in a −log_10_FDR of infinity. Detailed data can be found in Supplementary Table 5. **l:** Chord diagram showing GSEA enriched pathways in CESC, HNSC, HCC, LSCC, PDAC and UCEC. Supplementary Table 5. KIRC, kidney renal clear cell carcinoma; KICH, kidney chromophobe; KIRP, kidney renal papillary cell carcinoma; PRAD, prostate adenocarcinoma; CESC, cervical squamous cell carcinoma and endocervical adenocarcinoma; ccRCC, clear cell renal cell carcinoma; HNSCC, head and neck squamous cell carcinoma; LSCC, lung squamous cell carcinoma; UCEC, uterine corpus endometrial carcinoma; PDAC, pancreatic ductal adenocarcinoma. Statistical comparisons were made using the two-tailed Student’s t-test: *P < 0.05, **P < 0.01, ***P < 0.001.

## Discussion

Recognized as key pathogenic factors in RA-induced joint tissue destruction, RA FLSs have adopted distinct bioenergetic pathways that directly support their disease-inducing characteristics. Traits such as myofibroblast transformation, cytokine production, tissue invasiveness and pannus formation rely on alterations in fuel selection and energy carrier management ^45,46^. Cellular organelles, including mitochondria and the ER, are now understood to influence RA by deviating RA FLS transformation to tissue-destructive fibroblasts^47^. A significant effector pathway in RA involves the unchecked production of secretory proteins defect-targetable by anti-cytokine therapies. Our study suggests a hierarchical sequence with NAD^+^ metabolism malfunction leading to Golgi reshaping and protein secretion that further facilitates cell-cell interaction, thus exacerbating inflammatory responses^48^. This pathogenic pathway starts with suppressed QPRT, resulting in inadequate NAD^+^ production in the cytoplasm, ER and cis/medial Golgi and decreased global ADP-ribosylation and PARP12. The trans-Golgi network (TGN) detects this defect through suppressed ADP ribosylation and PARP12 expression. Suppressed ADP-ribosylation is proposed to induce the persistent activation of mTORc1 through inhibition of AMP-activated protein kinase (AMPK) nuclear release^26,27^, which might explain the increased mTORc1 activation in QPRT-suppressed cells. Hyperactivation of mTORc1 promotes the post-transcriptional regulation of EMT-associated secretome and phosphorylation of the medial-trans GRASP55. This phosphorylated GRASP55 promotes Golgi membrane unstacking, accelerating protein trafficking and vesicle formation^49^. Implicating NAD^+^ signaling in protein translation, Golgi integrity, and protein secretory function in RA pathogenesis adds a new layer to our understanding of autoimmunity and proposes novel treatment strategies for tissue inflammation, such as restoring QPRT activity or expression, replenishing NAD^+^ or restoring Golgi morphological defects.

Transfer of metabolic information to the secretory pathway relies on post-translational modifications of metabolic sensors localized in the organelles, thus allowing for rapid adaptation to environmental stimuli. For example, the generation of cytoplasmic NAD^+^ was recently proposed to govern the rough endoplasmic reticulum (rER) size through the ribosylation of the ER chaperone BiP, thus modulating TNF production in RA T cells^12^. NAD^+^ production, consumption and function are compartmentalized in the cell, underscoring the application of organelle-specific NAD^+^ biosensors for NAD^+^ metabolism and pathology studies. In this study, QPRT suppression depleted both cytoplasm and ER NAD^+^, accompanied by decreased ADP ribosylation and increased ribosome biogenesis. Therefore, it is tempting to speculate that increased protein production in QPRT^low^ RA FLSs might be partly due to expanded ER^12^ and mTORc1 translation control^38^, contributing to inflammation and RA FLSs aggressiveness. The current study further reveals that the QPRT suppression resulted in a global decrease in ADP ribosylation in RA FLSs. Under cellular stress, increased production and release of nuclear PAR leads to the relocation of PARP12 from the TGN to stress granules, resulting in mRNA translation blockade^50,51^. Interestingly, QPRT suppression was shown to induce PARP12 downregulation in RA FLSs, suggesting that lack of mRNA translation blockade due to insufficient PARP12 might be an alternative mechanism that promotes increased protein production in QPRT^low^ RA FLSs. Moreover, the inflated TGN-NAD^+^ following QPRT suppression might result from downregulated NAD^+^ consumption by the TGN-residing PARP12, making compartmentalized NAD^+^ an ideal indicator of Golgi dysfunction.

As the primary site for protein sorting and trafficking, the Golgi ultimately determines the fate of secretory proteins. We have identified the underlying defect in uncontrolled protein secretion in RA FLSs as a consequence of Golgi expansion, which increases the surface area for vesicle production and protein trafficking^40^. The dynamic structure and morphology of the Golgi are controlled by structural Golgi proteins, including Golgi reassembly and stacking proteins (GRASPs). The Golgi stacking and tethering regulator GRASP55 manages metabolic signals by controlling Golgi cisternae unstacking and unconventional protein secretion (UPS)^52^. Upon stress, inactivation of mTORc1 results in GRASP55 dephosphorylation, leading to its relocation from the Golgi to autophagosomes and multivesicular bodies (MVBs), where it facilitates autophagosome-lysosome fusion and UPS of surface and secretory factors^30^. Conversely, phosphorylated GRASP55 is proposed to localize in the Golgi and ER-Golgi interface^30,53^. In the Golgi, phosphorylated GRASP55 regulates protein trafficking by inducing Golgi fragmentation, possibly through disruption of the lateral connectivity of the Golgi ribbon and cisternae unstacking ^49^. Phosphorylated GRASP55 is also proposed to relocate to the ER-Golgi interface, inhibiting autophagy while aiding the Golgi bypass and secretion of specific secretory proteins ^53^. Data presented here demonstrated that the metabolic sensor GRASP55 interacts with PARP12, while depletion of QPRT inhibits this interaction. In QPRT^low^/PARP^low^ RA FLSs, GRASP55 deribosylation and phosphorylation is accompanied by persistent mTORc1 activity and Golgi ribbon unlinking. Functional analysis of QPRT-regulated genes consistently demonstrated enrichment of genes involved in the vesicle trafficking and ER-Golgi intermediate compartment. On the contrary, loss of QPRT or PARP12 disrupted GRASP55 interaction with autophagosomal LC3B and MVB-CHMP2A, suggesting that QPRT is required for the GRASP55-dependent UPS during stress. Data presented in this study proposes QPRT as a molecular switch that communicates metabolic dysfunction to the Golgi by regulating GRASP55-regulated Golgi-dependent and Golgi-independent protein secretion. It is worth noting that decreased NAD^+^ levels in the ER and cis Golgi might contribute to Golgi fragmentation through the decreased activity of sirtuins (SIRT) 2, which was proposed to deacetylate GRASP55, leading to its self-interaction^54^.

Directly targeting cytokines with biological therapies and small molecule inhibitors has transformed the management of multiple autoimmune diseases^55^. However, there remain unmet medical needs regarding therapeutic efficacy and safety profiles, as evidenced by limited disease remission rates and adverse effects^56^. Strategies that focus on reprograming hypersecretory pathogenic synovial cells into joint-protective/immunosuppressive cells could lead to novel tissue-protective immunomodulatory therapies. These therapies would address upstream pathologies rather than merely blocking the final product of proinflammatory and destructive effector cells. To explore new therapeutic strategies, we validated QPRT gene therapy interventions in an in vivo system of autoimmune arthritis. Golgi dysfunction and the resulting excessive cytokine production and joint tissue destruction were corrected by AAV-QPRT injection, suggesting that strategies to reprogram pathogenic protein production and secretion could focus on repairing the NAD^+^ metabolism and Golgi dysfunction. Metabolites or small molecules that stabilize and activate QPRT and/or PARP12 may be equally suitable for treating autoimmunity. QPRT modulation of pathological tissue-remodeling programs, such as EMT, suggests that shared metabolic mechanisms may regulate various cellular functions across different biological contexts. Illustratively, suppression of the PARP 12, identified in this study as a QPRT-regulated protein, is implicated in the dysregulated trafficking of E-cadherin to the plasma membrane and EMT-driven metastasis^43,57^. Therefore, targeting the QPRT-Golgi crosstalk could enable highly effective, pathogenesis-inspired therapeutic interventions for treating diseases in autoimmunity and beyond.

## Method

### Patients and samples

Human synovial tissue and blood samples were collected from individuals undergoing knee joint replacement surgery at the Shenzhen Second People’s Hospital in China. The participants with rheumatoid arthritis fulfilled the American College of Rheumatology and the European League Against Rheumatism diagnostic criteria and provided written informed consent^58^. The study was approved by the Ethics Committee of the Second People’s Hospital of Shenzhen (2023-157-01YJ).

### Cell isolation and culture

Rheumatoid arthritis synovial fibroblasts (RASFs) were isolated using the explant cell isolation method. Small synovial tissue explants were allowed to adhere to 75 cm^2^ culture flasks (Costar, Cambridge, MA, USA), whereafter fibroblast media (FM, Sciencell, 2301) supplemented with 1% penicillin/streptomycin (Gibco, 15140122) and 10% fetal bovine serum (FBS) (Gibco, 10099141). The explants were cultured at 37 °C in a humidified atmosphere of 5% CO_2_. Cells were grown to 80% confluence, detached with 0.25% trypsin-EDTA (Gibco, 25200072), split in a 1:3 ratio, and reseeded in FM under the same conditions. The cells used for our experiments were in passages 3 –7 because these cells were more homogenous than FLSs in the first and second passages and had better biological func tions than those older than the eighth passage.

### Cell lines

The immortalized RA-FLS MH7A cell lines (MH7A) were used for biosensor studies. These cell lines originated from RA synovium, as previously documented. The EA.hy 926 cell line, a human endothelial cell line, was used for tube formation. THP-1 is a human monocytic cell line derived from a patient with acute monocytic leukemia. MH7A and THP-1 cells were grown in RPMI 1640 medium (Gibco, C11875500BT) supplemented with 10% FBS, while EAhy926 were cultured in Dulbecco’s Modified Eagle Medium (DMEM, Gibco,11885)) supplemented with 10% FBS.

### Lentivirus packaging and cell transfection

The small hairpin RNA (shRNA) sequences were designed and synthesized by GenePharma (Shanghai, China). The sequences are listed in Supplementary Table 4. Non-targeting scrambled control (shCtrl) and QPRT-specific shRNAs in the pcDNA3.1+ vectors were packaged into lentiviral particles by cotransfecting HEK 293T cells with pMD2G (Addgene #12259) and psPAX2 (Addgene #394976) packaging vectors using polyethyleneimine (PEI, Yeasen, 40816ES03)transfection reagent. RA FLSs were plated in six-well plates at a density of 2 × 10^5^ per well. After 24 hours, the cells were infected with viral particles for 24 hours at 37°C with 8 μg/ml polybrene ( Yeasen, 40804es86). QPRT knockdown efficiency was confirmed by immunoblotting.

The biosensor coding sequences subcloned into the pCDH-CMV-MCS-EF1-Neo vector were previously developed in our lab^36^. The endoplasmic reticulum signaling sequence (signaling sequence of the calreticulin), cis/medial-Golgi (monoacylglycerol acyltransferases, MGAT 2) and trans-Golgi-targeting sequence (galactose-1-phosphate uridylyltransferase, GALT) were added to the N termini of the sensor. Using a PEI transfection reagent, the biosensor plasmids were packaged into lentivirus particles by cotransfecting 293T cells with pMD2G and psPAX2 packaging vectors. The immortalized RA-FLS (MH7A) were seeded in 60 mm Petri dishes at 80% confluency. After 24 hours, the cells were infected with viral particles for 24 hours at 37°C with 8 μg/ml polybrene. The transfected MH7A cells were then selected by serial dilution. Subcellular localization was confirmed by staining biosensor-expressing cells with Alexa Fluor 488 phalloidin (ThermoFisher, A12379), ER-Tracker Green (Beyotime, C1042M) and Golgi-Tracker Green (Beyotime, C1045S).

To evaluate the effect of QPRT knockdown on subcellular NAD^+^., MH7A cells stably expressing cytoplasm, ER, cis-Golgi and trans-Golgi biosensor were transfected with shQPRT lentivirus or shCtrl.

### Cellular BRET Measurement Using a Microplate Reader

The MH7A cell line that stably expresses the BRET sensor was seeded at a density of 10,000 cells per well in a white 96-well plate (Greiner) with DMEM culture supplemented with 10% FBS. After 24 hours of incubation at 37 °C with 5% CO_2_, the cells were starved for 12 hrs and then cultured in the absence or presence of recombinant human TNF-α (Peprotech, 300-01A). After 24 hours of treatment, the culture medium was aspirated and replaced with 100 μL of PBS buffer containing 1000 -fold diluted furimazine. Bioluminescence was measured using a FlexStation 3 Multi-Mode Microplate Reader (Molecular Devices) with NLuc emission measured at 440 nm and mScarlet-I emission at 590 nm or mNeonGreen emission at 515 nm.

### Condition medium preparation

QPRT or PARP12 knockdown RA FLSs were plated in a 10mm petri dish at 80% confluence. After 2 4 hours of incubation, the cells were washed with phosphate-buffered saline (PBS, Gibco, C10010500BT) and then incubated in DMEM containing 1% FBS for another 24 hours. The conditioned medium was collected by centrifugating the supernatant at 1200 rpm for 5 minutes to remove cell debris. An unconditioned 1% FM medium was used as a control.

### Matrigel in vitro tube formation assay

The in vitro tube formation assay assessed the impact of QPRT knockdown RA FLSs secretome on angiogenesis. Prechilled Angiogenesis µ-Slide 15 Well (Ibidi, Martinsried, Germany) plates were coated with 10µl Matrigel® Reduced Growth Factor Basement Membrane Matrix (Corning, 354234) and polymerized at 37°C for 30 minutes. 1 x 10^4^ human EA.hy926 endothelial cells in 50 μL conditioned or control medium without supplements were plated onto the pre-coated µ-Slide plates. The cells were incubated for 16 hours at 37°C, 5% CO_2_ atmosphere. Following incubation, cells were gently washed with HBSS and labeled with 8 μg/mL Calcein AM Fluorescent Dye (Corning, 354216) in HBSS (VivaCell, C3571-0500) for 30 minutes at 37°C, 5% CO2. After washing, images were acquired by fluorescent microscopy (Olympus) and capillary tube formation was quantified by averaging the number of tubes and branch points using the Angiogenesis analysis plugin of the Image J software. At least three fields per well were examined, with three replicates.

### Liquid chromatography-mass spectrometry (LC-MS) for metabolites

Blood samples were collected from RA patients and healthy volunteers, and plasma was obtained by centrifugation. The plasma was flash-frozen in liquid nitrogen and stored at –80°C. Plasma metabolites were extracted with 0.5 N perchloric acid solution, and the supernatant was filtered using a 0.45 μm PTFE syringe filter. Liquid chromatography-mass spectrometry (LC-MS) was conducted using the Agilent Infinity Lab LC/MSD iQ G6160A. The LC separation was achieved using a ZORBAX SB-Aq Column (Agilent, 4.6 × 100 mm, 3.5 μm). Solvent A consisted of 0.1% formic acid in water, while solvent B was acetonitrile. The gradient profile was as follows: 0 min, 20% A; 0.5 min, 20% A; 1.2 min, 60% A; 2.5 min, 60% A; 2.6 min, 20% A; 3 min, 20% A. Additional LC parameters included a flow rate of 1.00 mL/min, a column temperature of 20°C, and an injection volume of 5 μl. The mass spectrometer was equipped with an electrospray ionization (ESI) ion source and operated in positive ion mode for NAD ^+^ detection and in negative ion mode for KP metabolites (Trp, KYN, 3-HANA and QA). Metabolite data were analyzed using the Agilent OpenLab Data Analysis 2.205.7.2 software.

### Public bioinformatics dataset

To investigate the expression of QPRT in RA synovium, we analyzed human RA synovium datasets (GSE1919 and GSE89408) and mouse synovium datasets (GSE61140) obtained from Gene Expression Omnibus (GEO) using R (version 4.4.0). To investigate how QPRT influences RA synovium pathogenesis, data from five GEO datasets (GSE12021, GSE55457, GSE55584, GSE77298 and GSE89408), including 138 synovium samples from RA patients, were used to select samples with the top 17% highest and the top 17% lowest QPRT mRNA expression (23 samples in each group). The transcriptome data from these samples were analyzed using GSEA (version 4.2.3) to identify differentially expressed gene signatures. SRplot was used to visualize the volcano plot, heatmap and GO plots^59^.

The Cancer Genome Atlas (TCGA) datasets were preprocessed using R (version 4.4.0). Specifical ly, the TCGAbiolinks package was employed to load TCGA kidney renal clear cell carcinoma (TCGA_KIRC), TCGA prostate adenocarcinoma (TCGA_PRAD), TCGA kidney chromophobe (TCGA_KICH) and TCGA kidney renal papillary cell carcinoma (TCGA_KIRP) dataset. Gene expression data was then analyzed using the DESeq2 package to obtain log2-transformed counts. Clinical data were then integrated with gene expression data for downstream analysis. Differential expression analysis was performed using the limma package.

To download and preprocess clinical proteomic tumor analysis consortium (CPTAC) protein datasets, we utilized the CPTACBiolinks package in R. Available open-source data for clear cell renal cell carcinoma (ccRCC), head and neck squamous cell carcinoma (HNSCC), Lung squamous cell carcinoma (LSCC), uterine corpus endometrial carcinoma (UCEC) and pancreatic ductal adenocarcinoma (PDAC) proteomics data were obtained using getOmicsList(CancerType). Data normalization and quality control were conducted using GraphPad.

### Gene-set enrichment analysis

Gene-set enrichment analysis (GSEA 4.2.3) was performed to analyze datasets using 50 HALLMARK gene sets (v7.4). Genes were pre-ranked based on their RNA-seq expression counts and then ranked by GSEA software using the ‘Signal2Noise’ metric, with the permutation type set to ‘gene set’ and other default parameters (FDR threshold < 0.05). The HALLMARK gene sets enriched in QPRT^low^ tissues were ranked according to their enrichment scores.

### Immunoblotting

RA FLSs were harvested in radioimmunoprecipitation assay lysis buffer (RIPA; Solarbio, P0013B). Cellular proteins in the centrifuged supernatant were separated by 8 –12% sodium dodecylsulfate (SDS)–polyacrylamide gel electrophoresis (PAGE). Proteins on the gels were transferred to polyvinylidene fluoride (PVDF) microporous membranes (EMD Millipore, IPVH00010), which were then probed with various antibodies. The antibody company and dilution are presented in Supplementary Table 3. Tubulin expression served as the internal control.

### Quantitative real-time PCR assay

Total RNA was extracted with a Total RNA purification Kit (ES Science, RN001-100T) in RNase-free conditions according to the manufacturer’s instructions. cDNA synthesis was then performed using the Superscript First Strand Synthesis Kit (Invitrogen, Thermo Fisher Scientific, 11752 -250) according to the manufacturer’s instructions. Quantitative RT-PCR was performed with a SYBR Green PCR Master Mix (TaKaRa, RR820B) on the MxPro system. All primers were synthesized by the Qing Ke Biology (China). The primer sequences are listed in Supplementary Table 4. Gene expression was quantified relative to GAPDH using the 2^−ΔΔCT^ formula, and gene expression fold change was calculated relative to the control group.

### RNA sequencing

RNA-seq was conducted at the Beijing Genomics Institute (BGI). Each RNA sample included a mixture of RNA from three biological replicates. Library sequencing was performed with a 100-bp paired-end module. Raw fastq data were trimmed using Dr. Tom Multi-omics Data mining system (https://biosys.bgi.com) to generate clean fastq data and sequenced DNBSEQ high-throughput platform.

RNA-seq data were aligned to the hg19 genome and transcriptome using HISAT2 (v2.1.0) and aligned to the gene set using Bowtie2. The expression level of genes was calculated by RSEM (v1.3.1). Differential gene expression analysis was performed with DEseq2 at a threshold of FC > 1.5 and P < 0.05.

### LC-MS/MS analysis

LC-MS/MS analysis was conducted using a timsTOF Pro mass spectrometer (Bruker) coupled with a Nanoelute (Bruker Daltonics). Peptides were reconstituted with 40 µl of 0.1% (v/v) formic acid. Reverse-phase trap column (Thermo Scientific Acclaim PepMap100, 100 μm × 2 cm, nanoViper C18), self-packed with C-18 reverse-phase analytical column (Thermo Scientific Easy Column, 10 cm long, 75 μm inner diameter, 3 μm resin). The buffers used for chromatography were 0.1% formic acid (buffer A) and 84% acetonitrile / 0.1% formic acid (buffer B). Peptides were loaded onto the column at a 300 nL/min flow rate, controlled by IntelliFlow technology. The mass spectrometer was operated in positive ion mode. The mass spectrometer collected ion mobility MS spectra over a mass range of m/z 100-1700 and 1/k0 of 0.6 to 1.6 and then performed 10 PASEF MS/MS cycles with a target intensity of 1.5k and a threshold of 2500. Active exclusion was enabled with a release time of 0.4 minutes.

### MS data processing

Raw mass spectrometry data were processed using MaxQuant software (version 1.5.3.17). Database searches were conducted with the Andromeda search engine (version 1.5.3.17) integrated into MaxQuant against human UniProt databases. Default parameters in MaxQuant were used, except for the minimum ratio count and LFQ minimum ratio count, which were set to 1. Enzyme specificity was set to trypsin, allowing up to two missed cleavages. Cysteine carbamidomethylation © was set as a fixed modification, while oxidation (M) was set as a variable modification. The minimum peptide length required was 6 amino acids. Mass tolerances were set to 20 ppm. The “matching between runs” algorithm in MaxQuant was enabled. The false discovery rate (FDR) was estimated by searching against databases with reversed sequences, with a maximum FDR of 1% for protein and peptide identification.

Three independent biological replicates were performed. For protein quantification, LFQ intensity values from biological replicates, representing protein abundance, were used for statistical analysis. Protein identification requires at least two biological replicates and at least three peptides per protein.

### Proteomics data analysis

Hierarchical clustering was performed using Cluster 3.0 and Java Treeview, employing Euclidean distance and average linkage clustering, with heat maps and dendrograms for visualization. Motif analysis was conducted using MeMe, focusing on 13 amino acid sequences around the modified site. Subcellular localization predictions were made using CELLO, a multi-class SVM system. Protein domain signatures were identified with InterProScan, and Gene ontology (GO) annotations for biological processes were performed using NCBI BLAST+ and Blast2GO, plotting results by R scripts. KEGG annotations involved blasting proteins against the KEGG database to map pathways. Enrichment analysis was based on Fisher’s exact test with Benjamini-Hochberg correction, considering categories and pathways with p-values < 0.05 as significant. Protein-protein interaction networks were retrieved from IntAct and STRING databases, imported into Cytoscape for visualization, and analyzed to determine protein importance in the network.

### Cell invasion assay

The invasion assay was conducted using the Boyden chamber method with a transwell-65-mm insert, 8µm pore size polyethylene membrane (Transwell; Corning Labware Products). Briefly, 0.6ml FM containing 20% FBS was added to the lower chamber as the chemoattractant and 4.0 × 10^4^ FLSs suspended in 200 μl serum-free FM were seeded in inserts coated with 70 μl Matrigel. The culture plates were incubated for 24 h at 37 °C in a 5% CO_2_ atmosphere. The filters were then fixed with methanol for 10 min and stained with 0.1% crystal blue for 30 min. A cotton swab was used to remove cells from the upper surface of the membrane. Invaded cells were quantified using an inverted microscope (Olympus CKX53) and counted as the mean number of cells per 10 random fields in each assay.

### ELISA assay

- × 10^5^ cells were seeded into 12-well plates and cultured with FM without FBS. After 48 h, the supernatant was collected and centrifuged at 12000 rpm for 20 minutes. FGF2, VEGF and CTGF levels were measured using a Human FGF2, VEGF and CTGF ELISA kit (MultiScience) following the manufacturer’s protocols.

### Immunoprecipitation

HA-tagged GRASP55 or HIS-tagged PARP12 were transfected into RA FLSs. Cells were lysed in IP lysis buffer with protease, phosphatase, and protease inhibitors. The lysate was cleared by centrifugation, and immunoprecipitation was performed using anti-HA magnetic beads (HY-K0201, MedChemExpress) and anti-His magnetic beads (HY-K0209, MedChemExpress) at 4°C overnight. Complexes were washed, resuspended in 1×SDS sample loading buffer and analyzed by immunoblotting.

### Immunofluorescence

Cells were fixed with 4% paraformaldehyde for 10 min and permeabilized using 0.25% Triton X-100. The cells were incubated overnight with the respective antibodies: anti-GM130 (1:100, Immunoway), anti-GRASP55 (1:100, Proteintech) and PARP12 (1:100, Immunoway). After washing, the cells were then incubated with Cy3 goat anti-rabbit IgG (1:1000, Beyotime, catalog no. A0516) or Alexa Fluor-488 goat anti-rabbit (1:1000, Beyotime, catalog no. A0428) secondary antibody. Nuclei were stained with DAPI. The LSM710 system (Carl Zeiss) with a Plan Apochromat ×63/1.40 -NA oil DICIII objective lens (Carl Zeiss) was used to acquire images.

### Mice

The mice were housed under specific pathogen-free (SPF) mouse conditions at a constant room temperature of 22–26 °C and 40–60% humidity and free access to food and water under a 12 h light/dark cycle. Mice were randomly grouped according to weight. Experimental and control mice were co-housed in individual cages (maximum *n* = 5 per cage) while ensuring identical environmental conditions. The Animal Ethics Committee of the Chinese Academy of Sciences Shenzhen Institute of Advanced Technology (SIAT-IACUC-210309-YGS-CWX-A1756) reviewed and approved all animal procedures.

### AAV production

rAAV-CMV-WPRE-pA; AAV2/2 and rAAV-CMV-Qprt(m)-WPRE-pA; AAV2/2 vectors were production and quantification by BrainVTA. Briefly, HEK293T cells were at 50-70% confluence, cultured in 10 cm Petri dishes were triple transfected with pHelper, pAAV Rep-Cap, and pAAV ITR-expression plasmids in a 1:1:1 molar ratio using polyethyleneimine (PEI). After transfection, AAV was purified from the cell lysate and media using iodixanol density-gradient ultracentrifugation. Buffer exchange to 0.01% Pluronic P68 was performed using Amicon Ultra-100 kDa MWCO Centrifugal Filter Units. AAV titer was determined by SYBR Green qPCR with primers targeting the inverted terminal repeats within the AAV2 inverted terminal repeats. The vectors were stored at −80°C until use. The QPRT sequence is provided in Supplementary Table 4.

Eight-to ten-week-old male DBA/1J mice were obtained from Vital River (Guangdong Vital River Laboratory Animal Technology Co., Ltd.). To administer AAV, mice received an injection of 100µl of a viral solution containing 3×10^9^ viral genomes encoding either mouse QPRT (pAAV.QPRT) or empty vector (Empty.pAAV) in sterile saline. The injection was given intradermally, and the gene expression was confirmed by Western blot.

### Collagen-induced arthritis induction

Male DBA/1J mice were immunized with an emulsion consisting of 100 μg of chicken type II collagen (Chondrex, 20012) in an equal volume of 5 mg ml−1 complete Freund’s adjuvant (CFA) adjuvant (Chondrex, 7023) containing heat-killed Mycobacterium tuberculosis H37Ra (3.3 mg ml−1) administered intradermally at the base of the tail.

Clinical assessments were conducted once a week for 49 days and scored individually on a scale of 0 – 4, which resulted in a maximum score of 16. Each paw was scored as follows: 0, no evidence of inflammation; 1, swelling confined in a single digit; 2, swelling in tarsals and multiple digits; 3, moderate swelling extending from the ankle to metatarsal joints; 4, severe swelling encompassed the ankle, foot and digits, or ankylosis of the limb. The arthritic scores of four paws were summed.

### Micro-computed tomography scanning

At the end of the treatment period, the mice were euthanized, and the left hind paw of each mouse was amputated and fixed in 4% PFA. Three-dimensional micro-CT analysis was conducted on the left hind paws of healthy and arthritic mice. Scanning was performed using a Scanco VivaCT80 (SCANCO Medical, Switzerland) at 10.4 μm resolution with 70 kV tube voltage 114 μA tube current. The resulting three-dimensional microstructural image data were reconstructed, and structural indices were calculated using CT-Analyser (CTAn) software (Bruker). The Micro-CT scores were based on five disease-related indices: bone mineral density, bone volume fraction, cortical mineral density, trabecular number, and total porosity.

### Histological analysis

The hind limbs were fixed in 4% paraformaldehyde (PFA), decalcified in 10% ethylenediaminetetraacetic acid (EDTA, Aladdin, E116429), embedded in paraffin and sectioned at 5µM. Hematoxylin-Eosin (HE, Solarbio, G1120) staining was used to visualize inflammatory cell infiltration into the joints. Safranin O (Solarbio, G1371) staining assessed cartilage damage. Tartate-resistant acid phosphatase (TRAP, Solarbio, G1492) staining was performed to assess osteoclast activity. Histopathologic scores were calculated based on the ‘SMASH’ recommendations^60^. Data collection was performed in a blinded manner.

### Statistical analysis

Statistical significance was assessed using GraphPad Prism (v9.5.0, GraphPad Software) unless otherwise specified. The paired Wilcoxon or Mann-Whitney U test was utilized for quantitative data involving two groups with a sample size greater than five. Comparisons across three or more groups were conducted using one-way ANOVA followed by pairwise comparisons with Tukey’s method. For repeated measurements, such as arthritis scores, two-way ANOVA with post hoc testing using Bonferroni or Dunnett correction was employed to determine significance between groups. Proteomics data were initially analyzed using a two-tailed Student’s t-test, then calculating the false discovery rate (FDR) from the p-values and the number of tested variables using the Storey Tibshirani method for multiple hypothesis correction. All data are presented as mean ± s.e.m. *P < 0.05, **P < 0.01, ***P < 0.001. Detailed statistical parameters are presented in each figure legend.

## Data Availability

All data produced in the present study are available upon reasonable request to the authors

## ACKNOWLEDGEMENT

This work is supported by the National Key R&D Program of China (2021YFF1200300), National Natural Science Foundation of China (22207118, 22307135), Shenzhen Science and Technology Program (JCYJ20220818100804009), the Natural Science Foundation of Guangdong Province (2023A1515010715), R&D plan in key areas of Guangdong Province (2023B1111030002), Research Projects on Key Areas of General Colleges and Universities, Department of Education of Guangdong Province (2022ZDZX2071), Shenzhen Institute of Advanced Technology, Chinese Academy of Sciences.

## AUTHOR CONTRIBUTIONS

L.W.N., W.C., Q.Y., and K.L. conceived this study. L.W.N., K.L., C.X., L.C., P.W., G.H., Y.L., P.O.A., executed the experiments. Q.H. provided clinical samples. L.W.N., W.C., Q.Y., and K.L. performed public and experimental data analysis. All authors contributed to the manuscript writing.

## COMPETING INTERESTS

L.W.N. and Q.Y. are inventors of two patent applications filed by Shenzhen Institute of Advanced Technology. Patent application numbers are CN202310841862.6 and PCT/CN2023/137624.

---

**Extended Data Fig 1a:**
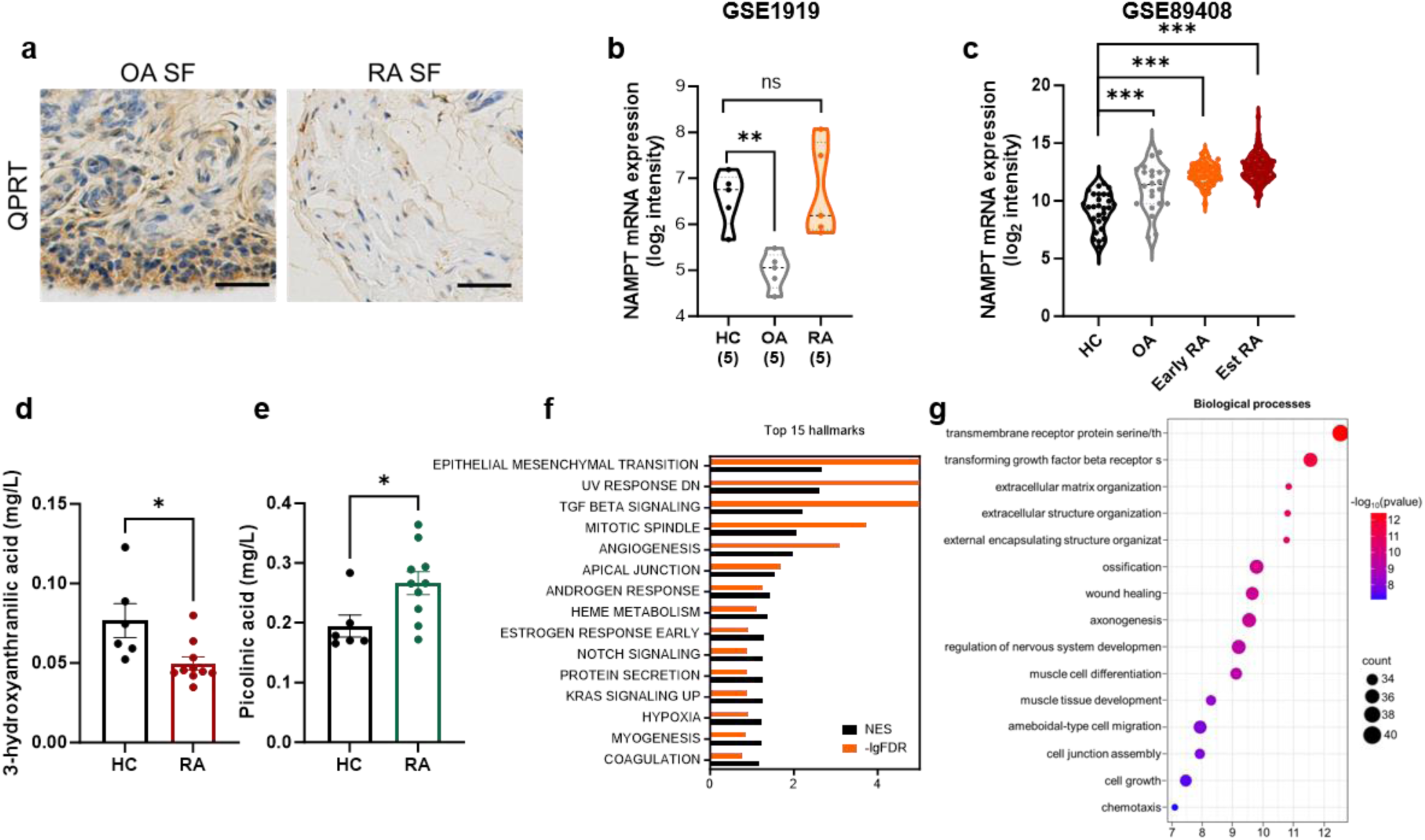
Immunohistochemistry staining of synovial tissue from OA and RA patients using antibodies against QPRT (brown) and counterstained with hematoxylin (blue). **b,c:** Comparison of NAMPT, rate-limiting enzyme in salvage de novo pathway, transcripts between healthy controls (HC) and RA human synovium from GSE1919 (a) and GSE89408 (b). **d-e:** LC-MS analysis of KP metabolites 3-hydroxy anthranilic acid and picolinic acid. **f:** Normalized Enrichment Scores (NES) with −log_10_FDR for enriched pathways identified in the GSEA analysis. The FDR is zero for the top three pathways, resulting in a −log_10_FDR of infinity. Detailed data can be found in Supplementary Table 1. **g:** Gene ontology (GO) biological processes of QPRT^low^ upregulated genes. The data are presented as the mean ± standard error of the mean (s.e.m.). ± s.e.m.; ns, not significant, *P < 0.05, **P < 0.01 and ***P < 0.001 compared to HC. Statistical significance was assessed by one-way ANOVA (**b,c**) or two-tailed Student’s t-test (**d,e**). The scale bars represent 100 μm. The enrichment bubble plot (**g**) was obtained from the SRplot web server^1^.

**Extended Data Fig 2a,b:**
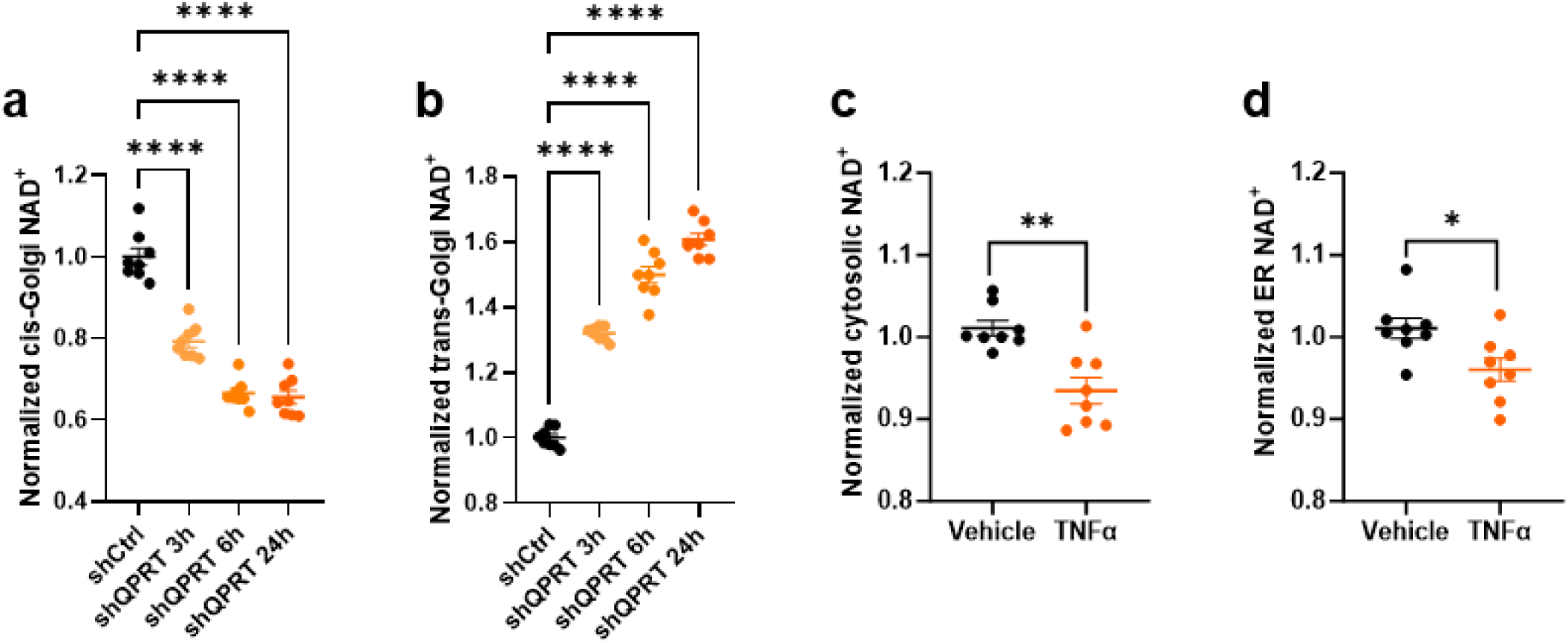
Time series quantification on the effect of shQPRT/shCtrl on cis/medial-Golgi and trans-Golgi. **c,d:** Quantification of the effect of TNFα (10ng/ml) on cytoplasmic and ER NAD^+^. Data are the mean ± s.e.m of independent replicates. P values were determined by one-way ANOVA (**a,b**) or two-tailed Student’s t-test (**c,d**): *P < 0.05, **P < 0.01 and ****P < 0.0001.

**Extended Data Fig 3a:**
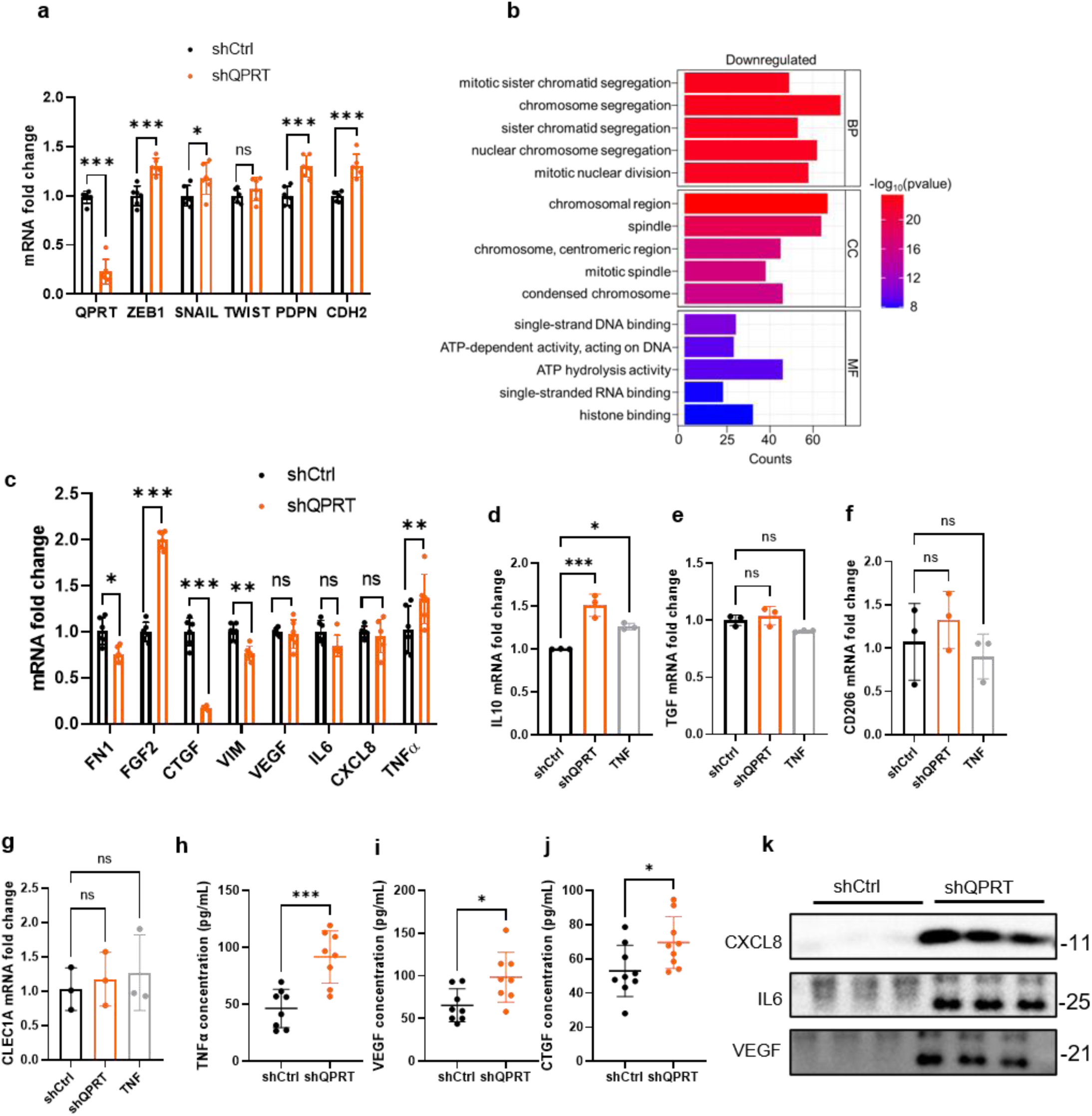
mRNA expression levels of QPRT, ZEB1, SNAIL, TWIST, PDPN and CDH2 in shQPRT/shCtrl transfected RA FLSs. **b:** Gene ontology (GO) analysis on downregulated genes. BP, biological properties; CC, cellular components; MF, molecular function. **c:** mRNA expression levels of EMT-related secretome **d-g:** mRNA expression levels of M2 macrophage markers IL-10, TGFβ, CD206 and CLEC1A in THP1 cell cultures in CM derived from shQPRT/shCtrl transfected RA FLSs. **h-j:** TNF (h), VEGF (i) and CTGF (j) levels in the supernatant were measured by ELISA. **k:** Representative immunoblots of CXCL8, IL6 and VEGF in the supernatant of shQPRT/shCtrl transfected RA FLSs. Data are mean ± s.e.m.; P values were determined by two-way ANOVA (**a,c**), one-way ANOVA (**d-g**) or two-tailed Student’s t-test (**h-j**): *P < 0.05, **P < 0.01, ***P < 0.001 and ****P < 0.0001.

**Extended data Fig 4a:**
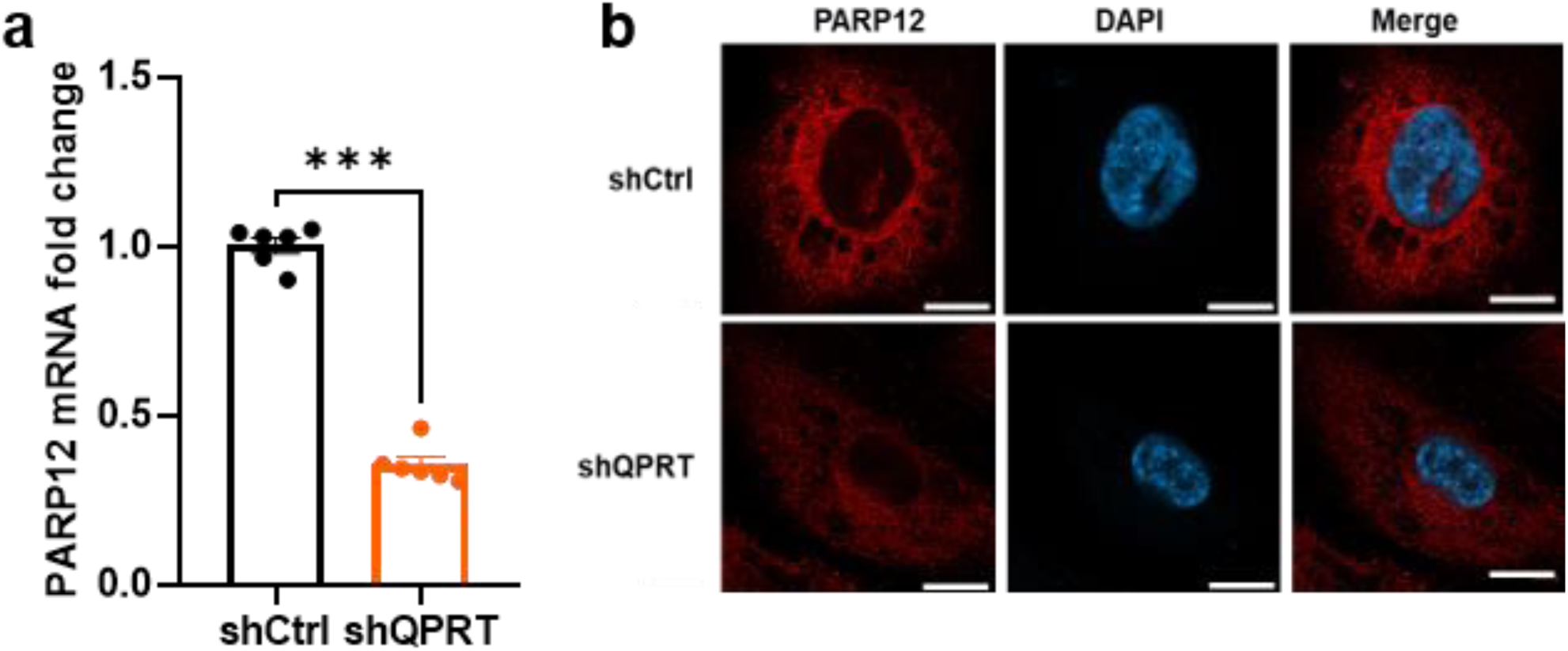
mRNA expression levels of PARP12 in shQPRT/shCtrl transfected RA FLSs. **b:** Representative immunofluorescence image of PARP12 expression in shQPRT/shCtrl transfected RA FLSs. **c:** Normalized Enrichment Scores (NES) with −log_10_FDR for significant pathways identified in the GSEA analysis are presented. Data are the means ± s.e.m and P values were determined by Student’s *t*-tests: \*\*\**P* < 0.001.

**Extended Data Fig 5a:**
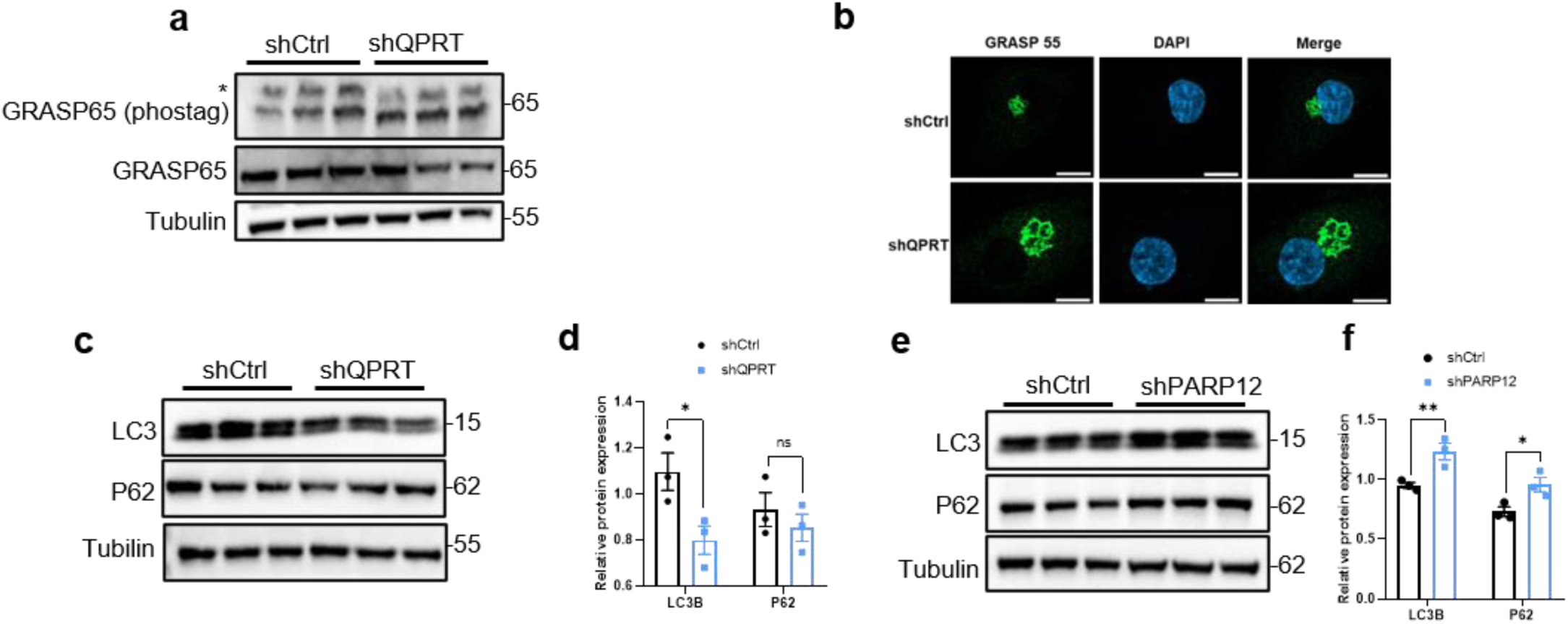
Immunoblotting analysis was performed on lysate from shCtrl or shQPRT transfected RA FLSs. GRASP65 phosphorylation was assessed using phos-tag gels, and the asterisk indicates p-GRASP65. **b:** Representative immunofluorescence image of GRASP55 expression in shQPRT/shCtrl transfected RA FLSs. **c-f:** Immunoblotting analysis of autophagy markers LC3B and P62 in shQPRT (c,d) or shPARP12 (e,f) transfected RA FLSs compared to control. Data are the means ± s.e.m. Statistical significance was assessed by two-way ANOVA (**d,f**): *P < 0.05, **P < 0.01, ***P < 0.001.

**Extended Data Fig 6a,b:**
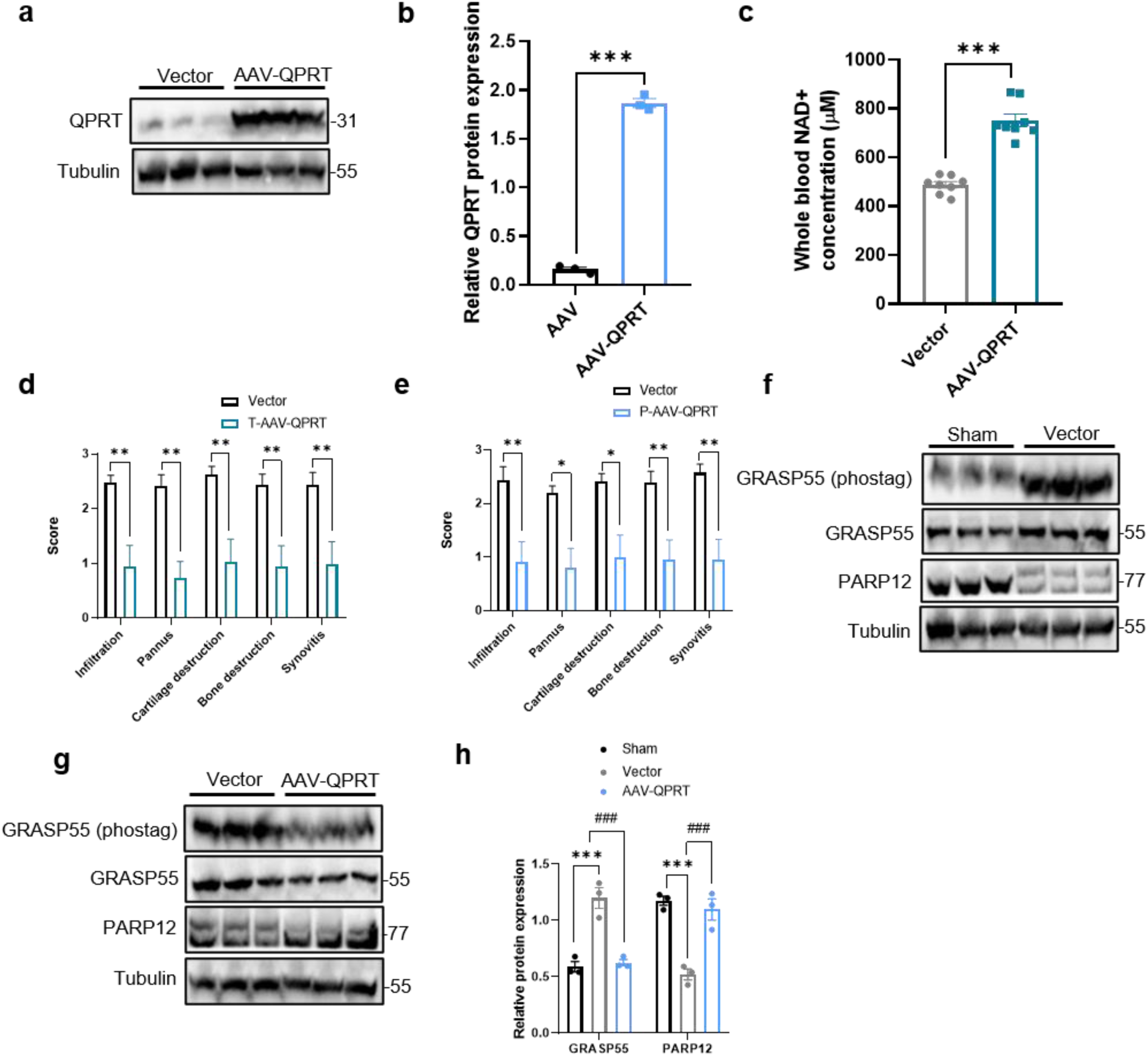
Immunoblotting analysis of QPRT expression in AAV and AAV-QPRT mice. Immunoblot represents data pooled from seven mice per group. **c:** Quantification of whole blood NAD+ from Vector and AAV-QPRT mice (n=8 per group). **d:** Pathological changes, cell infiltration, cartilage degradation, and bone destruction were assessed for the therapeutic regimen using the SMASH recommendations (Vector, n=7 and AAV-QPRT, n=8). **e:** Cell infiltration, cartilage destruction and bone destruction were evaluated for the prophylactic group ((Vector, n=7 and AAV-QPRT, n=8)). **f,g:** Hind paws from sham, Vector and AAV-QPRT were homogenized in cell lysis buffer for immunoblot detection of GRASP55 and PARP12. **h:** Quantification of GRASP55 and PARP12 levels in sham, Vector and AAV-QPRT mice. Immunoblot represents data pooled from eight mice per group. Statistical comparisons were made using two-tailed Student’s t-test (**b,c**) or two-way ANOVA (**d,e,h**): ***P < 0.001 compared to sham, ^###^P < 0.001 compared to Vector.

**Extended Data Fig 7a:**
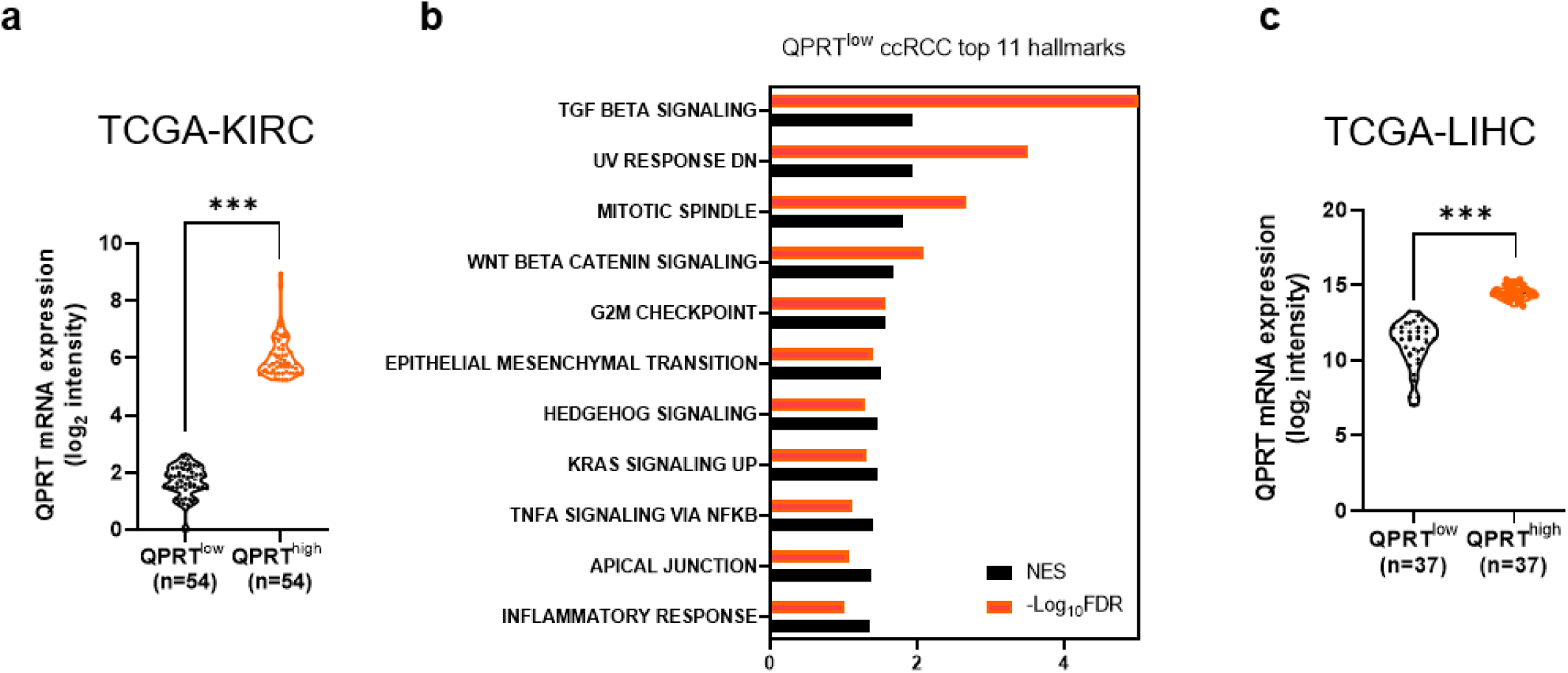
The top 10% QPRT^high^ expression samples (n = 54) and the bottom 10% QPRT^low^ expression samples (n = 54) from the cancer genome atlas (TCGA) kidney renal clear cell carcinoma (KIRC) samples. **b:** Normalized Enrichment Scores (NES) with −log_10_FDR for QPRT^low^ (TCGA-KIRC, n=54) enriched pathways identified in the GSEA analysis. The FDR is zero for the top three pathways, resulting in a −log10FDR of infinity. Detailed data can be found in Supplementary Table 5. **c:** The top 10% QPRThigh expression samples (n = 37) and the bottom 10% QPRTlow expression samples (n = 37) from the cancer genome atlas (TCGA) liver hepatocellular carcinoma (LIHC) samples.

## Notes

### Author Declarations

The study used openly available human data that were originally located at: Gene Expression Omnibus (GEO) The Cancer Genome Atlas (TCGA) clinical proteomic tumor analysis consortium (CPTAC) Human synovial tissue and blood samples were collected from individuals undergoing knee joint replacement surgery at the Shenzhen Second People's Hospital in China. The participants with rheumatoid arthritis fulfilled the American College of Rheumatology and the European League Against Rheumatism diagnostic criteria and provided written informed consent. The study was approved by the Ethics Committee of the Second People's Hospital of Shenzhen (2023-157-01YJ).

